# Phosphoproteomics uncovers exercise intensity-specific signaling networks underlying high-intensity interval training in human skeletal muscle

**DOI:** 10.1101/2024.07.11.24310302

**Authors:** Nolan J. Hoffman, Jamie Whitfield, Di Xiao, Bridget E. Radford, Veronika Suni, Ronnie Blazev, Pengyi Yang, Benjamin L. Parker, John A. Hawley

**Affiliations:** Exercise and Nutrition Research Program, Mary MacKillop Institute for Health Research, Australian Catholic University, Melbourne, Victoria, Australia; Computational Systems Biology Unit, Children’s Medical Research Institute, Faculty of Medicine and Health, The University of Sydney, Westmead, New South Wales, Australia; School of Mathematics and Statistics and Charles Perkins Centre, The University of Sydney, Sydney, New South Wales, Australia; Department of Anatomy and Physiology, The University of Melbourne, Parkville, Victoria, Australia; Centre for Muscle Research, The University of Melbourne, Parkville, Victoria, Australia

**Keywords:** HIIT, MICT, high-intensity interval training, moderate-intensity continuous training, exercise, skeletal muscle, kinase, signal transduction, phosphoproteome, mass spectrometry

## Abstract

In response to exercise, protein kinases and signaling networks are rapidly engaged in skeletal muscle to maintain energy homeostasis. High-intensity interval training (HIIT) induces superior or similar health-promoting skeletal muscle and whole-body adaptations compared to prolonged, moderate-intensity continuous training (MICT). However, the exercise intensity-specific signaling pathways underlying HIIT versus MICT are unknown. Ten healthy male participants completed bouts of work- and duration-matched HIIT and MICT cycling in randomized crossover trials. Mass spectrometry-based phosphoproteomic analysis of human muscle biopsies mapped acute signaling responses to HIIT and MICT, identifying 14,931 phosphopeptides and 8,509 phosphosites. Bioinformatics uncovered >1,000 phosphosites significantly regulated by HIIT and/or MICT, including 92 and 348 respective HIIT-specific phosphosites after 5 and 10 min and >3,000 total phosphosites significantly correlated with plasma lactate. This first human muscle HIIT signaling network map has revealed rapid exercise intensity-specific regulation of kinases, substrates and pathways that may contribute to HIIT’s unique health-promoting effects.

## INTRODUCTION

Exercise training confers numerous beneficial physiological adaptations in skeletal muscle, imparting a wide range of whole-body health benefits that can prevent, delay and/or treat a range of chronic metabolic conditions including obesity, type 2 diabetes and cardiovascular disease (Furrer et al., 2023a; Hawley et al., 2014). Protein kinases and downstream signal transduction networks mediate these adaptations and are activated/deactivated in contracting skeletal muscles in response to exercise in order to meet increased energy and oxygen demands and maintain cellular energy homeostasis (Egan and Zierath, 2013; Hargreaves and Spriet, 2020). Protein phosphorylation is a key post-translational modification that regulates nearly all cellular biological processes, including skeletal muscle adaptations to exercise. These exercise-induced adaptations in skeletal muscle are influenced by factors such as the mode, intensity, duration and frequency of exercise by engaging distinct molecular transducers (Hoffman, 2017).

High-intensity interval training (HIIT) has attracted widespread scientific and popular interest as a low volume, time-effective exercise intervention capable of inducing superior (Milanovic et al., 2015) or similar (Campbell et al., 2019) physiological adaptations (e.g., increased cardiorespiratory fitness) and reductions in cardiometabolic disease risk factors compared to higher volume, moderate-intensity continuous training (MICT). HIIT-based exercise protocols involve multiple (i.e., 4-10) work bouts of high-intensity exercise (≥ 80% v̇ O_2_max) interspersed with periods of rest or active recovery (Gibala et al., 2012; MacInnis and Gibala, 2017). A single bout of HIIT activates several key exercise-regulated kinases including the AMP-activated protein kinase (AMPK) and p38 mitogen-activated protein kinase (p38MAPK) in skeletal muscle, stimulating mitochondrial biogenesis and leading to increased mitochondrial content and enzyme activity in as little as 24 h post-exercise (Little et al., 2011; Perry et al., 2010). Relative to a workload-matched bout of MICT, a single bout of HIIT elicits similar skeletal muscle exercise-induced increases in AMPK signaling (Trewin et al., 2018), as well as p38MAPK and p53 tumor suppressor protein (p53) signaling responses that underpin mitochondrial biogenesis in human skeletal muscle (Bartlett et al., 2012). Other studies of HIIT using acute intermittent Wingate testing (i.e., four all-out 30 s exercise bouts interspersed with four min rest) report increased AMPK and p38 MAPK signaling activation immediately post-exercise (Gibala et al., 2009), but failed to detect any differences in signaling to key exercise-regulated kinases and gene expression responses involved in mitochondrial biogenesis compared to work-matched continuous exercise (Cochran et al., 2014). In contrast, increased activation of key signaling pathways including AMPK, p38 MAPK and Ca^2+^/calmodulin-dependent protein kinase II (CAMKII) has been detected in response to work-matched interval versus continuous exercise (Combes et al., 2015). However, in-depth analyses of the signaling networks engaged by HIIT versus MICT are lacking, with studies to date having analyzed only a subset of key exercise-regulated kinases, without investigating the breadth of potential signaling pathways underlying each exercise intervention.

Global mass spectrometry (MS)-based phosphoproteomics has the capability of identifying and quantifying thousands of protein phosphorylation events occurring within the complex and interconnected signaling networks engaged in response to exercise (Reisman et al., 2024). The first global phosphoproteomic analysis of exercise signaling in human skeletal muscle uncovered > 1,000 sites differentially phosphorylated after a short bout of continuous intense exercise (i.e., 8-12 min) versus rest (Hoffman et al., 2015). More recently, Blazev *et al*. utilized phosphoproteomics to map human skeletal muscle canonical signaling network responses to an acute bout of endurance (90 min cycling at 60% v̇ O_2_ max), sprint (cycling all out for 3 x 30 s) and resistance exercise (6 sets of 10 repetition maximum knee extensions) in eight healthy male participants, revealing 420 core phosphosites common to all exercise modalities (Blazev et al., 2022). This study also identified divergent signaling responses to these different modalities of exercise, although the exercise bouts were not matched for total workload nor duration. As such, it is difficult to isolate the divergent signaling responses related to exercise intensity, as kinase regulation can display different time profiles of activation/deactivation (Blazev et al., 2022). Furthermore, the common and unique kinases and downstream signaling pathways engaged by HIIT versus MICT in human skeletal muscle remain unexplored and no global phosphoproteomic analyses of HIIT-based exercise have been performed to date.

Therefore, the overall aim of this study was to determine the temporal regulation of kinases and downstream signaling pathways by an acute bout of HIIT versus MICT within the same human participants while controlling for total exercise duration, workload and diet. We hypothesized that HIIT would engage a unique subset of kinases and signaling pathways due to the intense, intermittent nature of this exercise modality, including increased activation of signaling networks underlying mitochondrial biogenesis. Utilizing a phosphoproteomic approach, this is the first study to map the human skeletal muscle signaling networks underlying an acute bout of HIIT and across different work-matched exercise intensities. We identify > 1,000 sites significantly regulated during (5 min) and immediately following (10 min) HIIT and/or MICT, including known and novel exercise-regulated signaling events. Furthermore, we identify a subset of kinases, substrates and pathways differentially regulated by HIIT relative to MICT and highly associated with plasma lactate responses to exercise, revealing the molecular framework underlying adaptive responses to HIIT that become rapidly engaged and contribute to HIIT’s potent ability to stimulate unique muscle physiological adaptations and whole-body health benefits.

## RESULTS

Ten healthy male participants were recruited and successfully completed all preliminary testing and the HIIT and MICT exercise bouts. The randomized crossover trial design permitted signaling responses to workload- and duration-matched HIIT and MICT exercise to be mapped and interrogated within the same participant (**Figure 1A**). Baseline whole-body anthropometric measurements and blood analyses confirmed these participants (age 25.4 ± 3.2 y; BMI 23.5 ± 1.6 kg/m^2^) were metabolically healthy, and maximal exercise capacity testing confirmed they were recreationally active but relatively untrained (relative peak oxygen uptake [v̇ O_2_peak] 37.9 ± 5.2 mL/kg/min) in line with our recruitment strategy to maximize detection of skeletal muscle signaling responses to exercise (**Figure 1B**). Peak power output (PPO; 208 ± 40 W) was used to prescribe the relative work-matched intensities for HIIT and MICT exercise. Lean mass from the DXA scan and resting metabolic rate (**Figure 1B**) were used to prescribe a standardized meal for each participant to consume prior to each exercise trial day.

**Figure 1.**
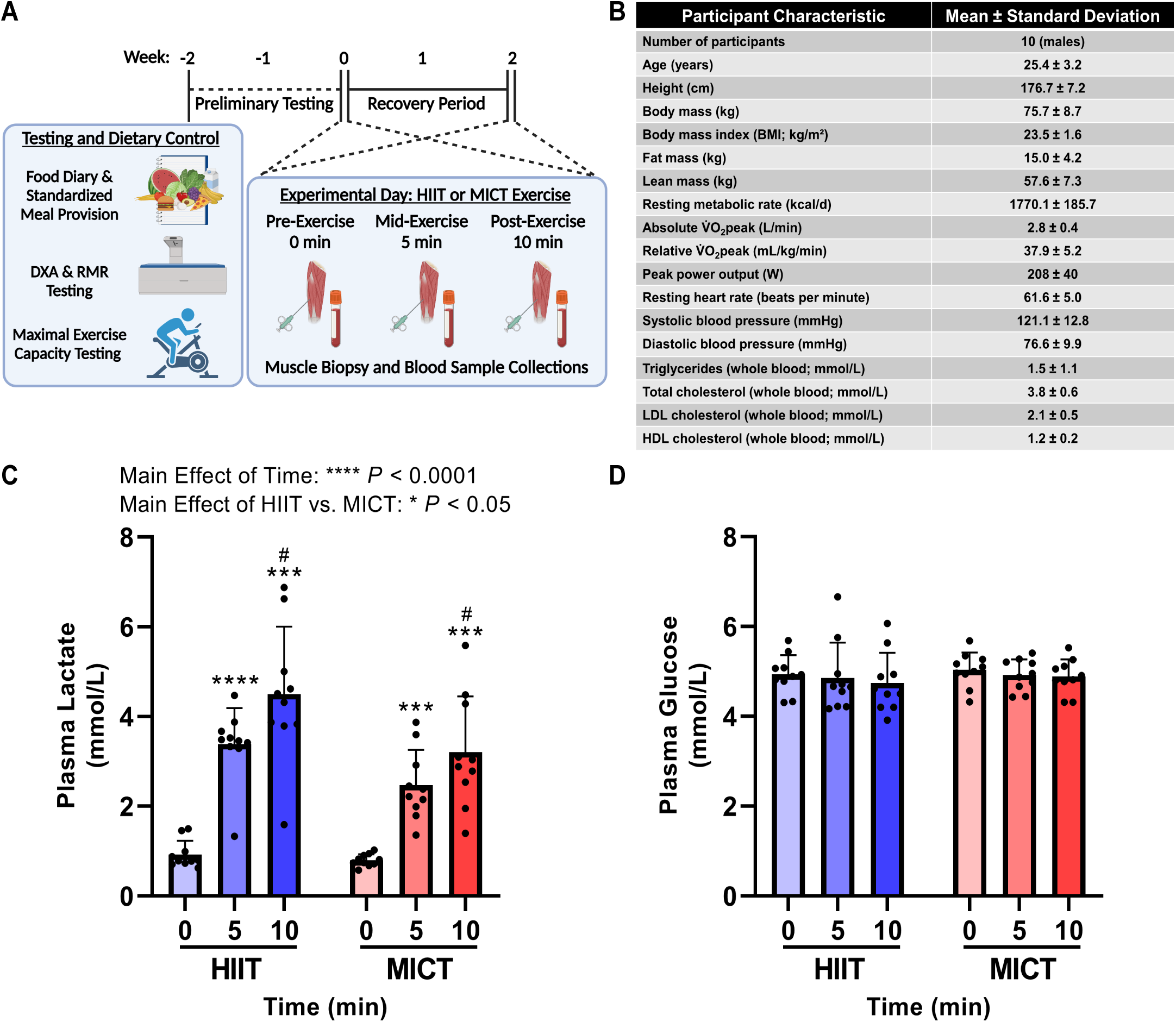
Preliminary testing and randomized crossover trial design, participant baseline characteristics, and plasma lactate and glucose responses to HIIT and MICT. As detailed in the overall study schematic (**A**), participants first underwent preliminary testing and dietary control prior to each experimental HIIT or MICT trial day. Participants arrived at the laboratory following overnight fasting for baseline measurements, a dual-energy x-ray absorptiometry (DXA) body composition scan and resting metabolic rate (RMR) testing. Each participant then completed an incremental fitness test to volitional fatigue on a cycle ergometer to determine peak oxygen uptake (v̇ O_2_peak) and peak power output (PPO) to calculate the work rate for the subsequent two workload (67.9 ± 10.2 kJ) and total duration (10 min) matched HIIT and MICT exercise trials. Participants’ food and fluid intake for all meals and snacks was recorded over a three-day period using a mobile phone application and analyzed by an accredited research dietician. A standardized dinner was consumed by each participant the evening prior to each exercise trial, with no caffeine or alcohol consumed 24 h prior. In a randomized crossover design, participants were randomly assigned their first exercise trial (i.e., HIIT or MICT) prior to commencing trial days and did not perform any exercise in the 72 h prior to each trial day. HIIT and MICT exercise trials were separated by at least a 10-day recovery period. On each experimental day, participants reported to the laboratory following overnight fasting, and vastus lateralis skeletal muscle biopsies and venous blood samples were collected pre-exercise (0 min), mid-exercise (5 min) and immediately post-exercise (10 min). Participant characteristics are listed in (**B**). Plasma (**C**) lactate and (**D**) glucose concentrations in response to HIIT versus MICT were determined using a YSI Analyzer. Data are presented as mean ± SD; two-way ANOVA with repeated measurements, Tukey’s test for multiple comparisons; *** *P* < 0.001 vs. 0 min; **** *P* < 0.0001 vs. 0 min; # *P* < 0.05 vs. 5 min; n=10 for each exercise intensity and timepoint.

Following consumption of a standardized dinner the evening before each trial, blood and skeletal muscle biopsy samples were collected in the fasted state at baseline and at 5 min and 10 min of each respective exercise bout (**Figure 1A**). Participants completing the acute bout of HIIT cycling, which consisted of 5 x 1-min work bouts at 85 ± 0.1% of individual PPO (176 ± 34 W) with 5 x 1-min active recovery intervals at 50 W, displayed increased plasma lactate concentrations at 5 and 10 min of exercise relative to pre-exercise baseline (**Figure 1C**; *P* < 0.0001 and *P* < 0.001, respectively; main effect of time *P* < 0.0001) with no changes in plasma glucose levels (**Figure 1D**). In response to a total work- and duration-matched acute bout of MICT (55 ± 2% individual PPO; 113 ± 17 W), participants displayed increased plasma lactate concentrations at 5 and 10 min of exercise compared to baseline (**Figure 1C**; *P* < 0.001; main effect of time *P* < 0.0001) with no changes in plasma glucose concentrations (**Figure 1D**). There was a main effect of higher plasma lactate levels in response to HIIT (**Figure 1C**; *P* < 0.05).

To map the signaling networks regulated by HIIT and MICT, proteins from each skeletal muscle biopsy sample were extracted, reduced/alkylated, digested to peptides, and isobarically labeled prior to phosphopeptide enrichment, fractionation and liquid chromatography with tandem MS (LC-MS/MS) phosphoproteomic analysis (**Figure 2A**). Principal component analysis (PCA; **Figure 2B**) and hierarchical clustering (**Figure 2C**) using the PhosR phosphoproteomic data analysis pipeline (Kim et al., 2021a) revealed a high level of consistency in the overall baseline signaling signature at rest (0 min) prior to the HIIT and MICT exercise trials, confirming our pre-exercise standardization strategies were effective. Distinct clusters were observed in response to the two divergent exercise challenges, with the 5 and 10 min HIIT signaling profiles clustering farthest away from the pre-exercise control profile (**Figure 2B-2C**). Overall, 19% of the total variance in the phosphoproteomic dataset was explained by principal component (PC)1, while PC2 explained 6% of the variance (**Figure 2B**). Global phosphoproteomics identified and quantified a total of 14,931 phosphopeptides (**Figure 2D, Supplemental Table 1**), corresponding to 8,509 phosphorylation sites (**Figure 2D, Supplemental Table 2**).

**Figure 2.**
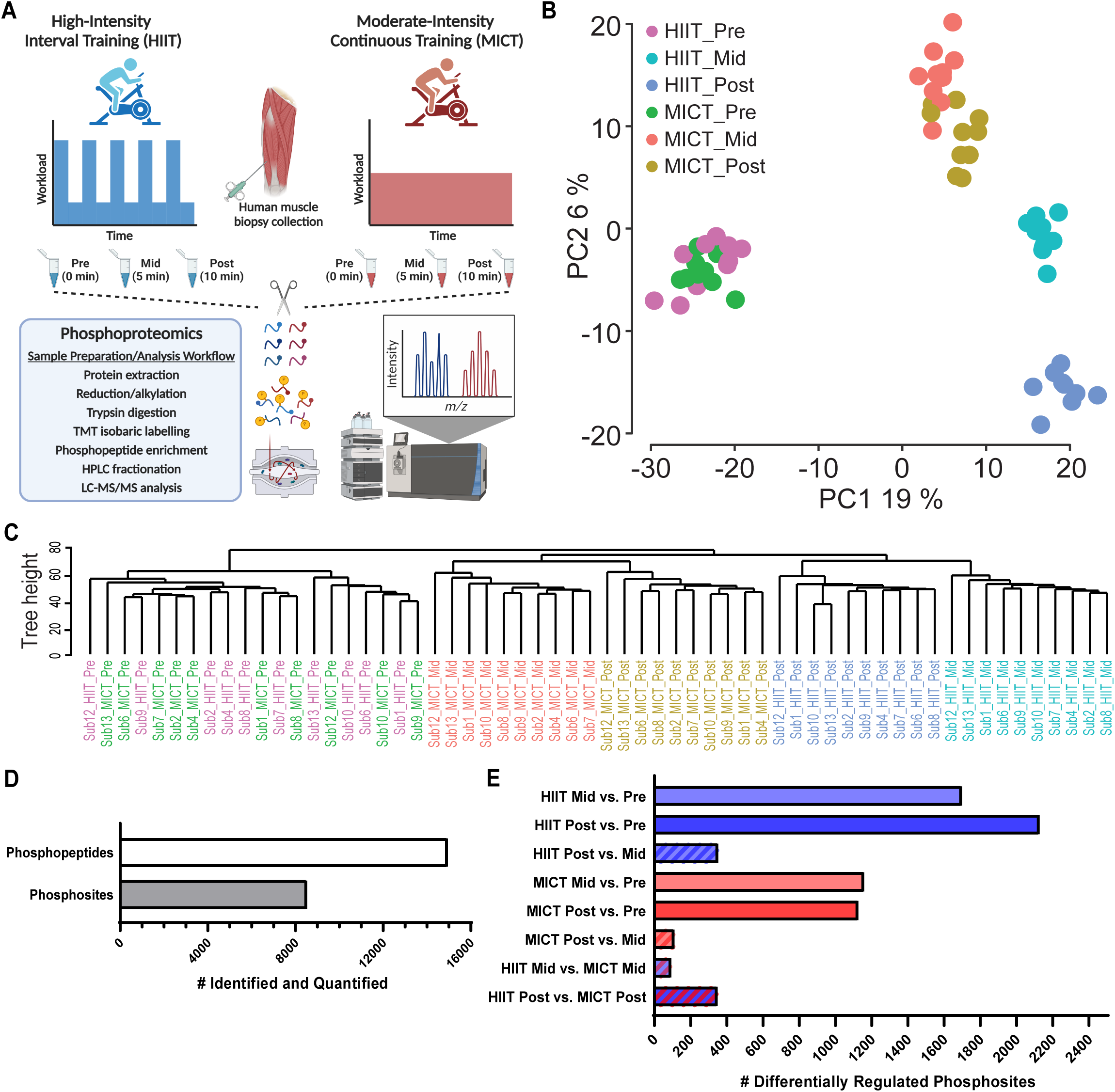
Human skeletal muscle phosphoproteomic analysis reveals effective pre-exercise standardization and distinct signaling profile clusters in response to HIIT versus MICT after 5 and 10 min. Vastus lateralis skeletal muscle biopsies were collected pre-exercise (0 min), mid-exercise (5 min) and immediately post-exercise (10 min) from each participant during the HIIT and MICT exercise trials (**A**). The 10-min HIIT cycling session consisted of alternating 1-min intervals at 85 ± 0.1% of individual Wmax (176 ± 34 W) and 1-min active recovery intervals at 50 W. The total duration- and work-matched MICT cycling session consisted of 10 min cycling at 55 ± 2% of individual Wmax (113 ± 17 W). Each of the 60 total muscle biopsy samples were prepared and subjected to LC-MS/MS analysis to accurately identify and quantity skeletal muscle protein phosphorylation sites at 0 min (pre-exercise), 5 min (mid-exercise) and 10 min (post-exercise) for both the HIIT and MICT exercise trials (**A**). Principal component analysis (**B**) and hierarchical clustering (**C**) of the phosphoproteomic datasets resulting from LC-MS/MS analysis of the 60 muscle biopsy samples was performed using the PhosR phosphoproteomic data analysis package (Kim *et al*. 2021 *Cell Reports*). Each individual point (**B**) or line (**C**) represents a unique biological sample, and samples are color-coded by exercise intensity and timepoint. Overall, 19% of the total variance in the overall phosphoproteomic dataset was explained by principal component (PC)1, while PC2 explained 6% of the variance. The total number of phosphopeptides and phosphosites identified and quantified using MS are shown (**D**), in addition to the number of differentially regulated phosphosites (-/+ 1.5-fold change and adjusted *P* < 0.05) from each timepoint and/or exercise intensity comparison (**E**).

Bioinformatic analyses of these phosphoproteomic data using PhosR (Kim et al., 2021a) identified >1,000 phosphosites significantly regulated (-/+ 1.5-fold change; adjusted *P* < 0.05; identified in ≥ 3 participants and ≥ 1 timepoint) by HIIT and/or MICT after 5 and 10 min, with < 400 of these sites differentially regulated post-versus mid-exercise (**Figure 2E; Supplemental Table 3**). Significantly regulated phosphosites observed in response to HIIT and/or MICT included kinases and substrates shown to be regulated by an acute bout of continuous high-intensity cycling such as those within the AMPK pathway (Hoffman et al. 2015; e.g., ACACB S222 (HIIT and MICT), AKAP1 S107 (MICT only), RPTOR S722 (HIIT and MICT), STBD1 S175 (HIIT and MICT; (Ducommun et al., 2019)), TBC1D1 S237 (HIIT and MICT) and TBC1D4 S704 (5-min HIIT only)). Multiple substrates of protein kinase A (PKA; PHKA1 S1018 and HSBP6 S16) were increased by both HIIT and MICT, while the PKA substrate CDK16 S110 was only increased after 10 min of HIIT. In addition, the known Akt substrate IRS1 S629 was only regulated after 10 min of both HIIT and MICT (Hoffman et al., 2015).

In response to HIIT, a total of 1,697 phosphosites (**Figure 3A**; including 1,235 and 462 down- and up-regulated, respectively) and 2,127 phosphosites (**Figure 3B**; including 1,580 and 547 down- and up-regulated, respectively) were significantly regulated after 5 and 10 min of exercise, respectively (**Figure 2E, Supplemental Table 3**). The top 10 sites displaying the most robust increases in phosphorylation in response to HIIT at 5 min versus rest included MYLPF T10, ZAC S454, FGA S285, MTFP1 S129, EEF2 T59, MAPRE2 S229, LMNA T424, HRC T207, ADSSL1 S7 and MTFP1 S128. The 10 phosphosites decreasing the most in response to HIIT at 5 min compared to rest included RTN4 S738, PDHA1 S262, DCLK1 S358, PDHA1 S201 and S269, SYNM T1109, MAP1B S1793, NGFR S217, SLC4A1 S349 and TRIP10 S354 (**Figure 3A; Supplemental Table 3**). After 10 min, top phosphosites most robustly increased by HIIT versus rest included CBX1 S89, MAPRE2 S229, MTFP1 S129, HRC T207, EEF2 T59, STBD1 S175, EEF2 T57, HRC S299, ZAK S454 and CIC S1371. Sites displaying the most decreased levels of phosphorylation by HIIT at 10 min included RTN4 S738, PDHA1 S201, SLC4A1 S349, KRI1 S142, NGFR S217, DCLK1 S358, CMYA5 S2825, TXNIP T294, CLASP2 S326 and SRRMS S2398 (**Figure 3B; Supplemental Table 3**). A total of 352 sites were differentially regulated after 10 versus 5 min HIIT (**Figure 2E**), including 212 down- and 140 up-regulated phosphosites (**Figure 3C; Supplemental Table 3**).

**Figure 3.**
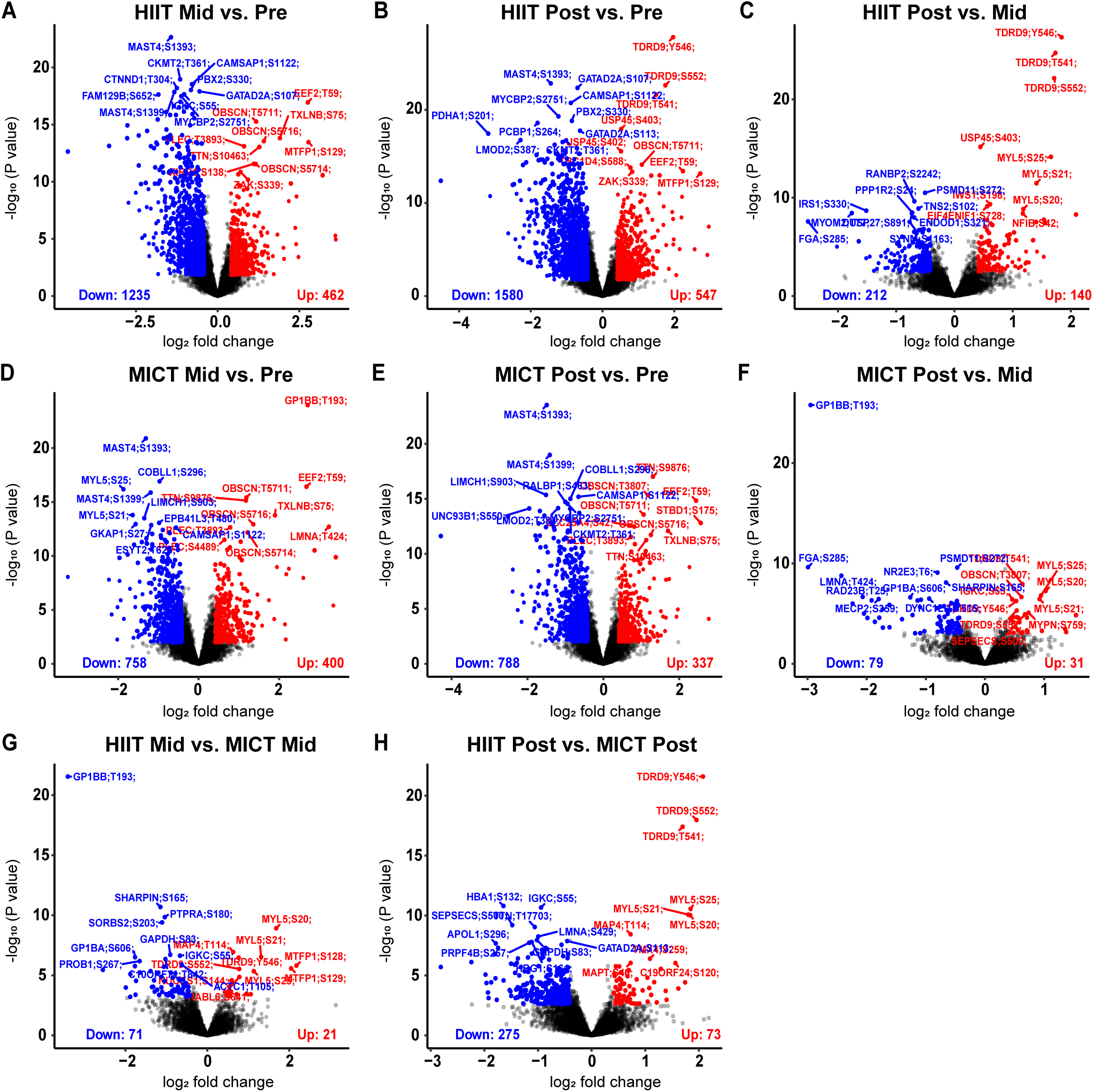
Human skeletal muscle phosphorylation sites differentially regulated by an acute bout of work- and duration-matched HIIT and/or MICT. Volcano plots showing the median phosphopeptide log_2_ fold change (x-axis) are plotted against the −log_10_ *P* value (y-axis) for each individual exercise intensity versus the respective timepoint (**A-F**). From the ∼15,000 total phosphopeptides detected, significantly up-regulated (red dots) and down-regulated (blue dots) phosphosites are shown (-/+ 1.5-fold change and adjusted *P* < 0.05), with black dots representing phosphosites that were detected but not significantly regulated by exercise. Volcano plots comparing signaling responses to each exercise intensity (i.e., HIIT versus MICT) at each timepoint are shown in **G-H**.

MICT induced changes in a lower number of significantly regulated phosphosites including a total of 1,158 phosphosites after 5 min (**Figure 2E and 3D**; 758 and 400 down- and up-regulated, respectively) and 1,125 phosphosites after 10 min (**Figure 2E and 3E**; 788 and 337 down- and up-regulated, respectively). A total of 110 sites were differentially regulated after 10 versus 5 min MICT, including 79 down- and 31 up-regulated phosphosites (**Figure 2E and 3F; Supplemental Table 3**).

To identify unique sites regulated by a single bout of HIIT relative to MICT, the signaling responses to each exercise intensity were next compared. After 5 min of HIIT, 71 phosphosites were down-regulated, while 21 phosphosites were up-regulated, relative to MICT (**Figure 2E and 3G; Supplemental Table 3**). These exercise intensity-specific signaling responses to HIIT were observed to be more robust at 10 min, with 275 phosphosites down-regulated and 73 phosphosites up-regulated compared to MICT (**Figure 2E and 3H; Supplemental Table 3**). Top exercise intensity-specific phosphosites most robustly up-regulated by HIIT relative to MICT included MTFP1 S128/S129 (increased by 5 and 10 min HIIT; S129 also increased by 5 and 10 min MICT), MYL5 S20/S21/S25 (increased by 10 min HIIT; decreased by 5 min MICT) and TDRD9 Y546/S552 (increased by 10 min HIIT; decreased by 5 min MICT). Exercise intensity-specific phosphosites down-regulated the most by HIIT versus MICT included GP1BB T193 (decreased by 5 and 10 min HIIT; increased by 5 min MICT), ATAD2B S374 (decreased by 5 and 10 min HIIT; decreased by 10 min MICT), MAPK1 T181 (decreased by 5 min HIIT; not regulated by MICT), ADD2 S532 and GP1BA S606 (decreased by 5 and 10 min HIIT; increased by 5 min MICT) and RYR1 T1399 (decreased by 5 and 10 min HIIT; decreased by 10 min MICT) (**Figure 3G-H; Supplemental Table 3**).

Kinase enrichment analyses were next performed to identify common and divergent activation/deactivation of kinases in response to the two exercise bouts (**Figure 4A**). Kinase activities were inferred via direction analysis using kinase perturbation analysis (kinasePA; (Yang et al., 2016)), to annotate and visualize how kinases with at least five quantified known substrates were perturbed by each exercise intensity and timepoint. These analyses confirmed activation/deactivation of known exercise-regulated kinases in response to both HIIT and MICT relative to pre-exercise, with significant kinase activity enrichments considered as kinases displaying | z-score | > 1.6. Kinases displaying similar increases in the levels of inferred kinase activity in response to both HIIT and MICT after 5 and/or 10 min included eukaryotic elongation factor 2 kinase (EEF2K), AKT1, AMPK alpha 1 catalytic subunit (PRKAA1), protein kinase cGMP-dependent 1 (PRKG1), and PKA subunit alpha (PRKACA), serum/glucocorticoid regulated kinase 1 (SGK1), ribosomal protein S6 kinase alpha-1 subunit (RPS6KA1), and MAP kinase-activated protein kinase 2 (MAPKAPK2). Moreover, exercise-induced decreases in the kinase activity of mammalian target of rapamycin (MTOR) were observed in response to both HIIT and MICT after 5 and 10 min. A strong trend for decreased kinase activity (i.e., z-score ∼ −1.6) of DNA-dependent protein kinase catalytic subunit (PRKDC) was also observed after 5 and/or 10 min of both HIIT and MICT.

**Figure 4.**
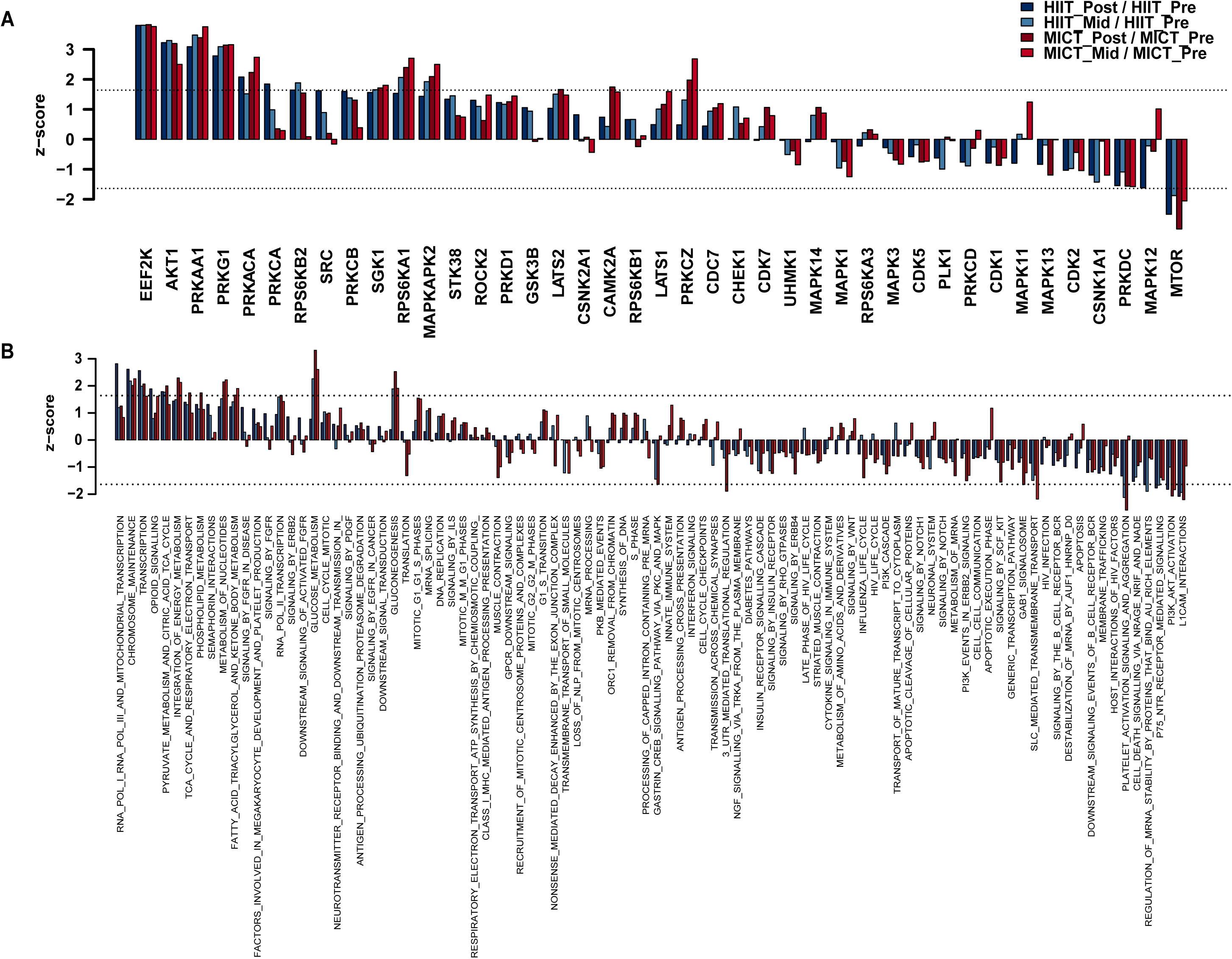
Kinase and pathway enrichment uncovers common and unique kinases and pathways regulated by HIIT and/or MICT. Kinase activity (**A**) was inferred via direction analysis using kinase perturbation analysis (KinasePA; (Yang et al., 2016)) to annotate and visualize how kinases and their known substrates are perturbed by each exercise intensity and timepoint. Pathway enrichment analysis (**B**) was performed using the Reactome database (Joshi-Tope et al., 2005) to determine biological pathways that are enriched within the lists of significantly regulated genes (corresponding to their respective phosphoproteins) for each exercise intensity and timepoint relative to its respective pre-exercise control. For kinase activity inference (**A**) and pathway enrichment (**B**), z-scores above and below the dotted lines (corresponding to | z score | > 1.6) were considered as increased or decreased by exercise, respectively.

Kinase enrichment analyses also revealed unique kinases that were differentially activated/deactivated (i.e., | z-score | > 1.6) in response to HIIT versus MICT after 5 and/or 10 min (**Figure 4A**). For example, activation of unique protein kinase C (PKC) conventional/atypical isoforms were observed to be differentially regulated by HIIT versus MICT, with increased activity of the conventional PKC alpha isoform (PRKCA; z-score > 1.6), as well as a strong trend for increased activity of the conventional PKC beta isoform (PRKCB; z-score ∼ 1.6), in response to only 10 min HIIT. In contrast, activity of the atypical PKC zeta isoform (PRKCZ) was increased only by 5 and 10 min MICT (z-score > 1.6). Other kinases shown to be uniquely activated by HIIT or MICT and/or displaying strong trends for exercise intensity-specific activation included S6 kinase beta-2 subunit (RPS6KB2; increased only by 5 and 10 min HIIT; z-score ≥ 1.6), proto-oncogene tyrosine-protein kinase SRC (only increased by 10 min HIIT; z-score ∼ 1.6), large tumor suppressor kinase 2 (LATS2; only increased by 10 min MICT; z-score ∼ 1.6), CAMK2A (increased only by 5 and 10 min MICT; z-score ≥ 1.6), large tumor suppressor kinase 1 (LATS1; only increased by 5 min MICT; z-score ∼ 1.6), and mitogen-activated protein kinase 12 (MAPK12; only decreased by 10 min HIIT; z-score ∼ −1.6).

Additional kinases that displayed unique and/or attenuated kinase activation trajectories in response to HIIT versus MICT but failed to reach significance in kinase enrichment analyses (**Figure 4A**; | z-score | < 1.6) included serine/threonine kinase 38 (STK38) with attenuated levels of activation in response to MICT relative to HIIT. In addition, glycogen synthase kinase 3 beta (GSK3B), casein kinase 2 alpha 1 (CSNK2A1), and ribosomal protein S6 kinase beta-1 subunit (RPS6KB1) all displayed trends for increased activation by HIIT with little or no activation detected in response to MICT. In contrast, polo-like kinase 1 (PLK1) and novel PKC delta isoform (PRKCD) showed trends for decreased activation in response to HIIT with little or no activation by MICT. Mitogen-activated protein kinase 11 (MAPK11) displayed opposite trends of activation between exercise intensities, with a tendency for decreased activation after 10 min of HIIT but increased activation after 5 min of MICT.

Pathway enrichment analyses were next performed using the Reactome database to determine biological pathways that were enriched within the lists of significantly regulated genes (corresponding to their respective phosphoproteins) for each exercise intensity and timepoint relative to pre-exercise, with significant enrichment considered as pathways displaying | z-score | > 1.6 (**Figure 4B**). Up-regulated Reactome pathways (i.e., z-score > 1.6) enriched in response to both HIIT and MICT after 5 and/or 10 min included expected exercise-regulated biological pathways such as ‘chromosome maintenance’, ‘transcription’, ‘opioid signaling’, ‘pyruvate metabolism and citric acid TCA cycle’, ‘glucose metabolism’ and ‘gluconeogenesis’. Down-regulated Reactome pathways (i.e., z-score < −1.6) enriched in response to both HIIT and MICT after 5 and/or 10 min included ‘L1CAM interactions’, ‘PI3K Akt activation’ and ‘platelet activation signaling and aggregation’.

The only Reactome pathway observed to be positively enriched in response to HIIT after 5 and/or 10 min but not enriched by MICT was ‘RNA polymerase I and III and mitochondrial transcription’ (**Figure 4B**). The only two pathways observed to be negatively enriched in response to HIIT after 5 and/or 10 min but not MICT included ‘P75 NTR receptor mediated signaling’ and ‘regulation of mRNA stability by proteins that bind AU rich elements’. In contrast, Reactome pathways positively enriched in response to MICT after 5 and/or 10 min but not enriched by HIIT included ‘integration of energy metabolism’, ‘TCA cycle and respiratory electron transport’, ‘phospholipid metabolism’, ‘metabolism of nucleotides’, and ‘fatty acid triacylglycerol and ketone body metabolism’. Pathways negatively enriched in response to MICT after 5 and/or 10 min that were not enriched by HIIT included ‘SLC mediated transmembrane transport’, ‘GAB1 signalosome’, ‘3’ UTR mediated translational regulation’, and ‘gastrin CREB signaling pathway via PKC and MAPK’.

Kinase-substrate associations were next predicted in response to HIIT and MICT via the phosphoproteomic signaling profiles and kinase recognition motif of known substrates using kinasePA (**Figure 5**; (Yang et al., 2016)). This analysis generated predictions matrices, with columns corresponding to kinases, rows corresponding to phosphosites, and values denoting how likely a phosphosite is phosphorylated by a given kinase in response to HIIT (**Figure 5A**) and MICT (**Figure 5B**). Pathway enrichment analyses were then performed using phosphosites with a high prediction score for each kinase to observe how biological pathways containing substrate phosphosites are commonly and/or uniquely regulated by HIIT (**Figure 5C**) versus MICT (**Figure 5D**). Collectively, these analyses revealed that a range of kinases displayed unique patterns and/or significance levels of signaling pathway regulation in response to each exercise intensity including HIPK2, GSK3B, CDK6, CDK2, CDK1, CDK7, MAPK14, MAPK3, CHEK1, PRKCG, CDK5, PRKDC, PIM2, PRKCQ, PRKCD, CSNK2A1, PLK1, AKT1, PRKCB and PRKCA (**Figure 5C-D**).

**Figure 5.**
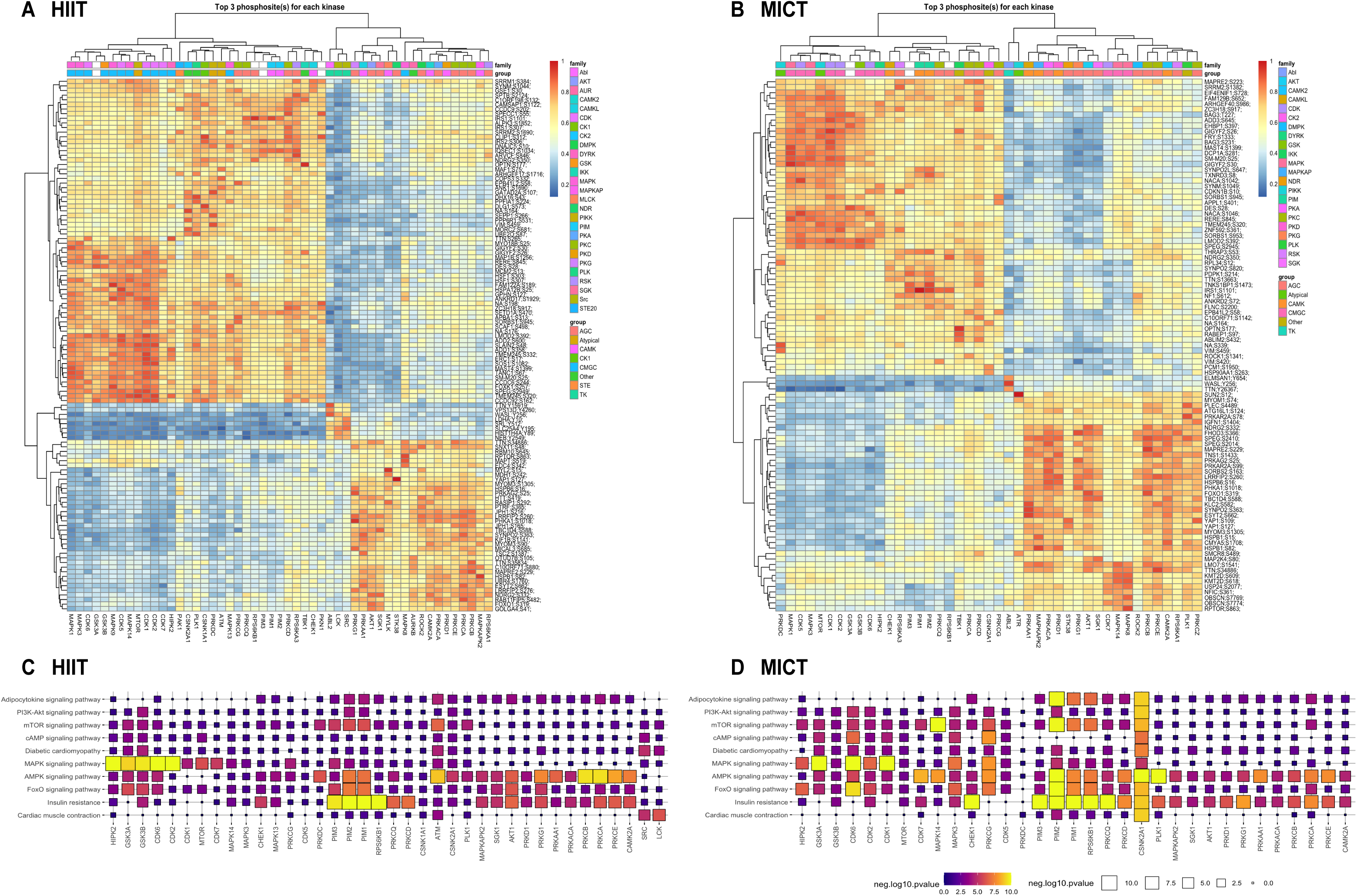
Kinase-substrate predictions and pathway enrichment analysis identify differential regulation of downstream substrates and pathways in response to HIIT versus MICT. Kinase-substrate associations were predicted in response to HIIT and MICT via the phosphoproteome signaling profiles and kinase recognition motif of known substrates using PhosR (Kim et al., 2021a). This analysis generated predictions matrices, with columns corresponding to kinases, rows corresponding to phosphosites, and values in the heat maps denoting how likely a phosphosite is phosphorylated by a given kinase in response to HIIT (**A**) and MICT (**B**). Pathway enrichment analysis was performed using kinase-substrate predictions (i.e., phosphosites with a high prediction score for each kinase) to determine how kinases regulate common and/or distinct signaling pathways in response to HIIT (**C**) and MICT (**D**).

Using these matrices, signalome networks were then reconstructed based on the correlation between kinases to visualize how kinases and substrates are regulated in response to HIIT (**Figure 6A**) versus MICT (**Figure 6B**). This revealed distinct signalome profiles between HIIT and MICT, with kinases clustered closer together more highly correlated in terms of their kinase-substrate predictions. To determine the proportion of common and unique substrates targeted by specific kinases in response to HIIT versus MICT, phosphoproteins were clustered into protein modules according to their prediction matrices shown in **Figure 5A-B**. This clustering revealed five unique clusters of HIIT and MICT kinase-substrate signaling profiles (**Supplemental Figure 1A-E**; modules 1 to 5). Visualization of these five clusters for individual kinases revealed shared or unique substrate signaling trajectories in response to HIIT (**Figure 6C**) versus MICT (**Figure 6D**) within their unique signalome networks. For example, widely studied exercise-regulated kinases including AKT1, CAMK2A, MAPK1, MAPK3, MTOR and PRKAA1 displayed similar substrate clusters between exercise intensities, while other kinases such as ABL2, MAPK14, PLK1, PRKCA, PRKCE, RPS6KA1 and STK38 regulated unique substrate clusters in response to HIIT versus MICT (**Figure 6C-D**).

**Figure 6.**
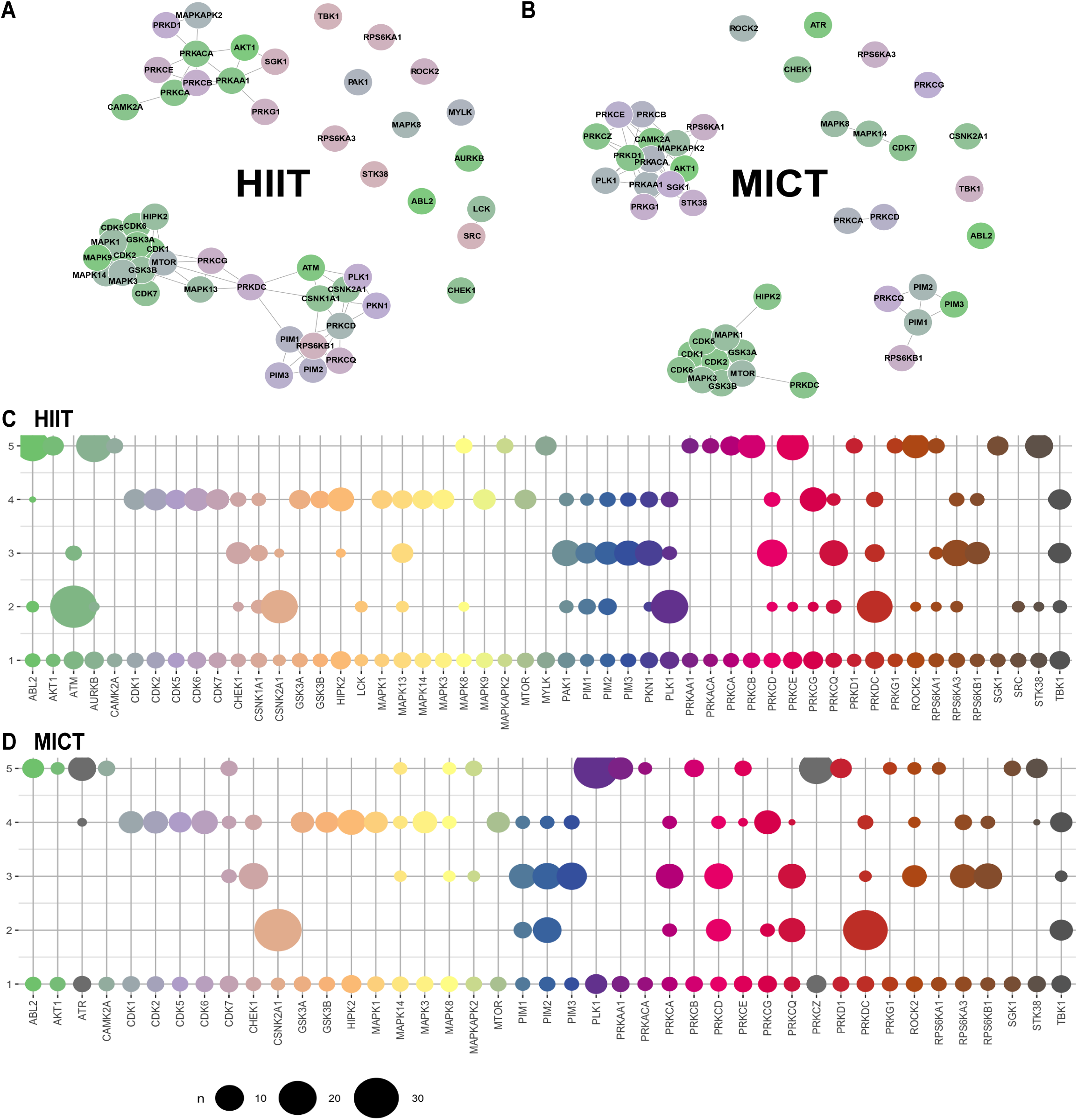
Signalome network highlights distinct HIIT and MICT kinase clusters and differential signaling trajectories in response to each exercise intensity. Signalome networks for HIIT (**A**) and MICT (**B**) exercise were constructed using the PhosR phosphoproteomic data analysis package (Kim et al., 2021a). This ‘signalome’ construction method utilized the phosphoproteome signaling profile and kinase recognition motif of known substrates to visualize the interaction of kinases and their collective actions on signal transduction. Kinases clustered together are highly correlated in terms of kinase-substrate predictions. Visualization of five phosphoprotein clusters from the phosphoproteomic dataset highlights distinct kinase-substrate regulation within the HIIT and MICT signaling networks, with shared and unique signaling trajectories shown for a panel of kinases in response to HIIT (**C**) and MICT (**D**).

Finally, to demonstrate the utility of the global phosphoproteomic dataset and test the hypothesis that higher plasma lactate concentrations during HIIT can influence which skeletal muscle exercise-regulated signaling proteins become engaged, correlation analyses were performed to interrogate individual phosphorylation site responses within the dataset relative to the plasma lactate responses to HIIT and MICT exercise. A total of 3,084 phosphosites were significantly correlated with plasma lactate concentrations from each timepoint and exercise intensity (**Supplemental Table 4**; q < 0.05 with Benjamini Hochberg false discovery rate (FDR)). Among these phosphosites, nine of the top 50 most significantly correlated phosphosites have annotated functional roles in regulating their respective protein’s activation state in the PhosphoSitePlus annotation database (**Figure 7A-D**; (Hornbeck et al., 2012)). For example, four of these top nine phosphosites that were significantly correlated with plasma lactate have experimentally validated roles in governing their respective protein’s activity and are involved in regulating a range of key metabolic processes (e.g., glycolysis, glucose transport and mitochondrial biogenesis) including PDHA1 S201 (**Figure 7A**), RPTOR S859 (**Figure 7B**), TFEB S123/S128/S136 (**Figure 7C**) and TBC1D4 S588 (**Figure 7D**). Other phosphosites among the top 10% most significantly correlated sites with plasma lactate concentrations included known AMPK-regulated sites (e.g., ACACB S222; mitochondrial fission regulator-1 like protein (MTFR1L) S141 (Tilokani et al., 2022)), as well as novel phosphosites with no known functional role (e.g., DENND4C S989, phosphoprotein present in glucose transporter GLUT4 vesicles; MTFP1 S128/S129, phosphoprotein involved in inner mitochondrial membrane fission/fusion) that may respectively be involved in regulating skeletal muscle glucose transport and mitochondrial network dynamics in response to exercise.

**Figure 7.**
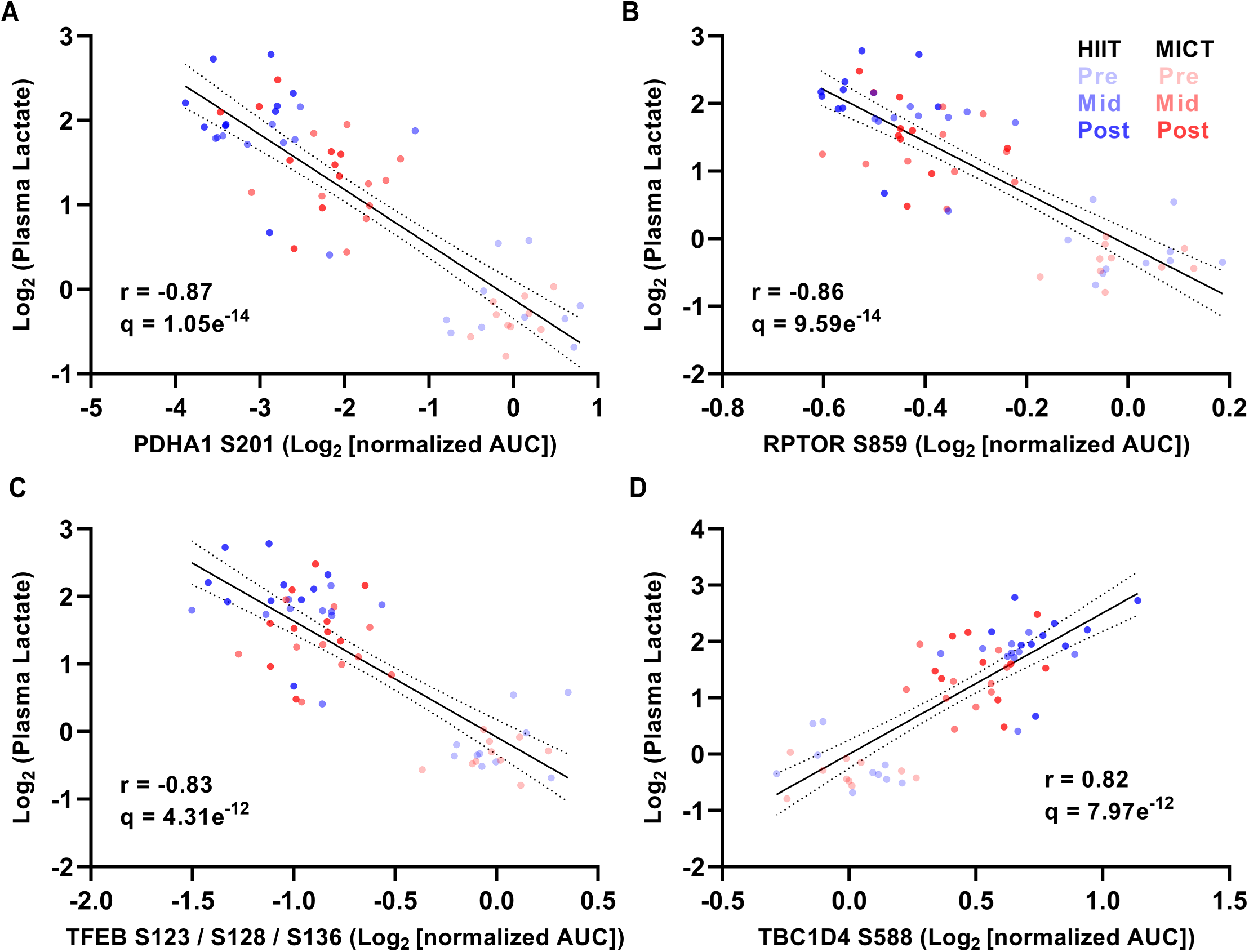
Correlation of HIIT and MICT phosphosites and plasma lactate levels identifies > 3,000 lactate-correlated sites including functional phosphosites that govern protein activity and metabolic regulation. Spearman correlation of individual phosphorylation sites (n=8,509 total) with plasma lactate concentrations at each timepoint and exercise intensity (n=60 total plasma samples analyzed) are shown for four of the most significantly correlated sites (q < 0.05 with Benjamini Hochberg FDR) with annotated functional roles in governing their respective phosphoprotein’s activation state and regulating a range of key metabolic processes (e.g., glycolysis, glucose transport and mitochondrial biogenesis) including PDHA1 S201 (**A**), RPTOR S859 (**B**), TFEB S123/S128/S136 (**C**) and TBC1D4 S588 (**D**).

## DISCUSSION

For the first time, this analysis of the HIIT signaling network in human skeletal muscle has revealed the early time course of acute signaling events underlying muscle adaptive responses to HIIT versus MICT. Our global phosphoproteomic approach has revealed the rapid induction of a complex network of common and exercise intensity-specific kinases, substrates and pathways regulated by a single bout of HIIT versus work-matched MICT after just 5 min of exercise, that were highly correlated with the prevailing plasma lactate responses to each exercise intensity. To the best of our knowledge, this is the first phosphoproteomic study of exercise signaling that has matched for workload and duration to assess divergent signaling responses between exercise intensities.

The matching of total workload and duration between exercise bouts is important and contrasts previous phosphoproteomic studies that have compared exercise signaling responses to divergent workloads of differing durations and modalities (Blazev et al., 2022). Such an approach permitted resolution of time-sensitive profiles of distinct signaling responses comparing the effects of a moderate-intensity continuous exercise challenge to a high-intensity interval-based exercise protocol within the same participants in a crossover study design, allowing sufficient recovery (i.e., > 10 days) to ensure no residual effects of the previous exercise bout. Standardized meal provision before each trial prescribed in accordance with each participant’s body composition, resting metabolic rate and food diary records led to a highly reproducible resting ‘baseline’ signaling signature for each participant, confirming a high level of control and reinforcing the importance of utilizing dietary standardization in addition to just overnight fasting to help control for potential differences in nutrient/energy availability between trials.

This study utilized a metabolically healthy, but untrained participant cohort to induce robust signaling responses, as available evidence indicates exercise-induced signaling responses in skeletal muscle such as AMPK activation are more pronounced in untrained relative to trained human participants (McConell et al., 2005; Yu et al., 2003). All ten participants were able to successfully complete both work-matched exercise trials, with the HIIT protocol eliciting higher plasma lactate concentrations compared to MICT. The two exercise intensities both provided a robust contractile stimulus that engaged a range of known exercise-regulated kinases including activation of AMPK (regulated by AMP/ATP and ADP/ATP ratios), EEF2K (regulated by Ca^2+^/calmodulin), PKA (regulated by cAMP) and PRKG1 (regulated by NO/cGMP). Activation of AKT1 increased after 10 min (e.g., inferred via known substrate IRS1 S629), while surrogate markers of MTOR activation decreased (e.g., NDRG2 T248 decreased only after 10 min HIIT; NDRG2 S344 increased only after 5 and 10 min MICT), in response to both exercise intensities. Overall, a higher number of phosphosites were down-regulated versus up-regulated at each timepoint and exercise intensity, suggestive of acute exercise-regulated kinases becoming inhibited and/or phosphatases becoming activated early after commencing exercise.

In this study mitochondrial fission process 1 (MTFP1; also known as MTP18) S128/S129 were identified among the top HIIT-regulated sites, with S128 only regulated by HIIT and more robust phosphorylation of S129 observed following HIIT versus MICT. The inner mitochondrial membrane protein MTFP1 has known roles in mitochondrial fission/fusion and was previously identified as having increased abundance in rat diaphragm following endurance exercise training (Sollanek et al., 2017), but has not been implicated in acute exercise signaling to date. MTFP1 has recently emerged as a key regulator of liver mitochondrial and metabolic activity, with liver-specific deletion of MTFP1 in mice conferring protection against high fat diet-induced metabolic dysfunction and hepatosteatosis via upregulating oxidative phosphorylation activity and mitochondrial respiration (Patitucci et al., 2023). MTFP1 has been identified as a gene target of the master regulator of skeletal muscle mitochondrial biogenesis, the peroxisome proliferator-activated receptor gamma coactivator 1-alpha (PGC-1α), in C2C12 myotubes (Nsiah-Sefaa et al., 2014) with its translation regulated by mTORC1 activity (Morita et al., 2017). Human skeletal muscle MTFP1 gene expression, concomitant with PGC-1α expression, was recently shown to be reduced by leg immobilization and increased upon resumption of physical activity and resistance exercise training (Pileggi et al., 2023). Collectively, this growing body of evidence suggests that unique phosphorylation of MTFP1 in response to HIIT may represent a novel mechanism underlying exercise-regulated mitochondrial inner membrane fission and/or fusion. Further investigation of MTFP1’s potential functional roles in maintaining skeletal muscle mitochondrial networks and metabolic homeostasis is warranted.

The observed differences in work-matched HIIT versus MICT signaling responses may be due to stochastic changes or accumulation of intracellular calcium and/or other metabolites in response to fluctuating energy demands during HIIT’s repeated work-rest cycles (Gibala and Hawley, 2017), and is consistent with other studies investigating effects of acute work-matched bouts of HIIT versus MICT (Bartlett et al., 2012; Peake et al., 2014). Indeed, investigations that have determined metabolic fluctuations occurring in work-matched interval versus continuous exercise have observed a greater level of activation of kinases such as AMPK, CaMKII and p38 MAPK, suggesting that buildup of upstream stimuli for these kinases (e.g., increased AMP/ATP and/or ADP/ATP ratios, intracellular calcium) may trigger greater activation in response to interval-based exercise (Combes et al., 2015). While levels of AMPK activation were similar between HIIT and MICT in this study, differential activation of PKC conventional/atypical isoforms suggests potential influence of muscle intracellular calcium during HIIT. For example, activation of the atypical PKC isoform PRKCZ was only observed in response to 5 and 10 min MICT, supporting previous observations of human muscle PRKCZ phosphorylation and/or activation in response to continuous cycling at ∼70-75% v̇ O_2_peak (Perrini et al., 2004; Rose et al., 2004), as rapidly as the first 5 min after commencing exercise (Rose et al., 2004). The present study has revealed previously unappreciated exercise intensity-specific activation of calcium-responsive conventional PKC isoforms (i.e., PRKCA, and a trend for increased activation of PRKCB) in response to HIIT, suggesting intracellular calcium spikes during HIIT intervals may drive PKC isoform-specific activation. As no further increase in atypical PKC isoform activity was observed in previous studies in response to increased continuous exercise intensity (Richter et al., 2004), these data suggest the stochastic nature of HIIT may govern its unique activation of PKC isoforms versus MICT. Furthermore, activation of the calcium-responsive CAMK2A, which is rapidly activated in human muscle in response to continuous exercise (Rose and Hargreaves, 2003; Rose et al., 2006), was differentially regulated by HIIT versus MICT (i.e., CAMK2A activity only increased by 5 min MICT). This suggests differential signaling mechanisms underlying calcium homeostasis during continuous versus interval exercise may contribute to HIIT’s potent ability to stimulate mitochondrial biogenesis (Torma et al., 2019).

Rates of carbohydrate oxidation are increased, while rates of fat oxidation are reduced, in response to HIIT compared to MICT (Peake et al., 2014). HIIT increases levels of skeletal muscle glucose transport, glycogenolysis and glycolysis, leading to accumulation of glycolytic products such as muscle lactate that can act as signaling molecules and influence exercise-regulated signaling networks within skeletal muscle, as well as be released into the bloodstream to facilitate interorgan communication with other tissues such as the heart, liver and brain (Brooks et al., 2023; Hargreaves and Spriet, 2020). Plasma lactate responses to HIIT in the present study were increased relative to MICT, with > 3000 phosphosites identified as being significantly correlated with the prevailing plasma lactate concentrations. Among the most highly correlated phosphosites with plasma lactate were PDHA1 S201 (**Figure 7A**) and TBC1D4 S588 (**Figure 7D**) with annotated functional roles in the regulation of glycolysis and glucose transport, respectively. PDHA1 is a subunit of the PDH enzyme complex, which is regulated by pyruvate and ADP and becomes activated by calcium during exercise, serving as a key link between glycolysis and the tricarboxylic acid (TCA) cycle by converting pyruvate to acetyl-CoA (Hargreaves and Spriet, 2020).

TBC1D4 is a key nexus in skeletal muscle glucose transport regulation via controlling glucose transporter GLUT4 translocation in response to insulin and muscle contraction during exercise (Cartee, 2015). Given the strong associations (i.e., | r | > 0.80) observed between plasma lactate concentrations and these key regulatory phosphosites in the present study, and in light of other studies investigating signaling-metabolite correlations in response to distinct exercise modalities and workloads (Blazev et al., 2022), these correlation data support the hypothesis that lactate accumulation in skeletal muscle and/or blood is a major driver of exercise intensity-specific physiological adaptations (MacInnis and Gibala, 2017). Collectively, differences in muscle and circulating metabolites and counterregulatory hormones in response to HIIT versus MICT may therefore influence metabolite-protein interactions during/after 10 min of exercise, leading to exercise intensity-specific signaling events and unique physiological adaptations. In addition to increasing plasma lactate concentrations, the higher exercise intensity of HIIT versus MICT may also lead to differential muscle fiber type recruitment that in turn influences fiber type-specific signaling pathway responses such as AMPK (Kristensen et al., 2015), which were undetectable given the present study’s focus on analyzing whole muscle biopsy samples.

There are several limitations of this first global study of the HIIT signaling network that warrant further investigation in future studies of human skeletal muscle exercise signaling. First, this study involved only male participants to allow us to benchmark observed signaling responses to previous studies investigating continuous exercise protocols in males. The acute responses of skeletal muscle to HIIT in female human participants remain uncharacterized and are being actively investigated in our ongoing research. The HIIT and MICT protocols utilized were selected based on the feasibility of untrained participants to complete each exercise bout. As a result, 5-10 min of HIIT or MICT at the prescribed intensities may not have engaged the full repertoire of shared and exercise intensity-specific signaling pathways in muscle and may require longer exercise durations in future studies. For example, patterns of substrate oxidation change during more prolonged exercise and differ between sexes, which may affect signaling responses more robustly between exercise intensities and/or sexes.

The signaling responses and programmed muscle adaptations to an acute bout of HIIT or MICT may also not be able to predict chronic training adaptations (Hawley et al., 2018). For example, recent transcriptomic and proteomic analyses of mammalian skeletal muscle have highlighted that acute changes may not be predictive of chronic exercise training-induced changes (Furrer et al., 2023b), such as HIIT-induced remodeling of the skeletal muscle proteome after repeated training (Hostrup et al., 2022; Robinson et al., 2017). Extensive total proteome analysis was not performed in this study, as only 5-10 min of exercise has been previously observed to not affect total muscle protein content (Hoffman et al., 2015). While not analyzed, given the acute nature of the exercise bouts in this study, repeated HIIT training (i.e., six sessions per week for two weeks) can stimulate more robust mitochondrial adaptations (i.e., greater exercise-induced increases in citrate synthase maximal activity and mitochondrial respiration) compared to total work and duration-matched MICT in human skeletal muscle, suggesting that exercise intensity and/or the pattern of muscle contraction may drive peripheral adaptations to exercise (MacInnis et al., 2017). The amplification of human skeletal muscle signaling and mRNA responses underlying mitochondrial biogenesis in response to acute low-volume, high-intensity exercise may be due to more robust induction of metabolic stress relative to more prolonged, moderate-intensity exercise (Fiorenza et al., 2018; MacInnis and Gibala, 2017). In this regard, it has been speculated that accumulation of skeletal muscle intracellular AMP, ADP, calcium and/or other metabolites such as lactate during HIIT intervals may differentially affect skeletal muscle signaling responses and adaptations to exercise such as mitochondrial biogenesis relative to continuous bouts of exercise (MacInnis et al., 2019). However, with repeated chronic HIIT versus MICT training, exercise training volume may become more important than intensity in increasing skeletal muscle mitochondrial content over time (Bishop et al., 2019).

Finally, the functional relevance of HIIT and/or MICT regulation of unique kinase activation patterns including isoform-specific PKC activation and novel HIIT-regulated phosphosites such as MTFP1 S128/S129 remains unknown. Functional validation experiments (e.g., target knockdown, overexpression and/or mutagenesis) in cellular and/or animal models are required to determine their respective roles in regulating exercise-induced muscle adaptations and molecular processes, such as MTFP1’s potential functional role in regulating inner mitochondrial membrane fission/fusion in response to acute exercise. Regardless of these considerations, the present study has uncovered a previously unknown breadth of shared and exercise intensity-specific molecular machinery engaged by a single bout of HIIT versus MICT in untrained healthy males. Unique exercise signaling pathways and molecular transducers uncovered within these HIIT and MICT signaling networks can ultimately be leveraged and reinforced with longer interventions and/or repeated exercise training to stimulate cardiometabolic health-promoting effects and help combat a range of chronic metabolic conditions in populations with disease.

## Data Availability

This article and its Supplemental Information include all datasets generated during this study. The MS phosphoproteomics data have been deposited to the ProteomeXchange Consortium via the PRIDE (Perez-Riverol et al., 2022) partner repository with the dataset identifier PXD053295 (Reviewer login details - Username; Password: 1PKEot15EUSc). Further information and requests for materials and resources including raw data, code and unique materials collected and used in this study should be directed to and will be fulfilled by the corresponding author and lead contact, Nolan J. Hoffman.

https://www.ebi.ac.uk/pride

## ACKNOWLEDGMENTS

This work was supported by Australian Catholic University (ACU) research funding awarded to N.J.H. N.J.H. and J.A.H.’s research is partially funded by the Australian Government through the Australian Research Council (ARC) Discovery Project grant DP200103542, ‘Molecular networks underlying exercise-induced mitochondrial biogenesis in humans’. B.L.P. is funded by an Australian National Health and Medical Research Council (NHMRC) Emerging Leader Investigator Grant (APP2009642). We are grateful to all participants for their time and dedication to this study. We thank Andrew Garnham for performing muscle biopsy and blood sample collections, Rebecca Hall for dietary assistance, and Natalie Janzen, Mehdi Belhaj, Imre Kouw, Evelyn Parr and Marcus Callahan for laboratory support on trial days. Figures 1A and 2A were created with BioRender.com.

## AUTHOR CONTRIBUTIONS

N.J.H. and J.A.H. conceptualized the study and provided intellectual input and financial support. N.J.H., J.W. and B.E.R. conducted participant baseline measurements, exercise trials and sample collection. N.J.H., J.W. and B.E.R. performed skeletal muscle, blood and whole-body physiological data analysis. R.B. and B.L.P. performed MS sample preparation and phosphoproteomic analysis. D.X., V.S. and P.Y. performed bioinformatic analyses. N.J.H. wrote the manuscript, and all authors edited and approved the final version.

## RESOURCE AVAILABILITY

This article and its Supplemental Information include all datasets generated during this study. The MS phosphoproteomics data have been deposited to the ProteomeXchange Consortium via the PRIDE (Perez-Riverol et al., 2022) partner repository with the dataset identifier PXD053295 (Reviewer login details – Username; Password: 1PKEot15EUSc). Further information and requests for materials and resources including raw data, code and unique materials collected and used in this study should be directed to and will be fulfilled by the corresponding author and lead contact, Nolan J. Hoffman.

## SUPPLEMENTAL INFORMATION TITLES AND LEGENDS

**Supplemental Figure 1.** Protein modules showing distinct clusters of kinase-substrate signaling profiles in response to HIIT versus MICT.

**Supplemental Table 1.** Human skeletal muscle phosphopeptides identified and quantified using TMT labeling and following Perseus data processing.

**Supplemental Table 2.** Human skeletal muscle phosphosites identified and quantified following PhosR data normalization.

**Supplemental Table 3.** Human skeletal muscle differentially regulated phosphosites from each timepoint and exercise intensity.

**Supplemental Table 4.** Phosphosites significantly correlated with plasma lactate concentrations from each timepoint and exercise intensity.

## METHODS

### Ethical approval

This study was approved by the Australian Catholic University Human Research Ethics Committee (approval number 2017-311H), prospectively registered with the Australian New Zealand Clinical Trials Registry (registration number ACTRN12619000819123) and conformed to the standards set by the Declaration of Helsinki. All participants completed medical history screening to ensure they were free from illness and injury and were informed of all experimental procedures and possible risks associated with this study prior to providing their informed consent.

### Human participants

Ten healthy males aged 18-30 yr with a body mass index 18.5 – 27.0 kg/m^2^ were recruited to participate in this study. Participant characteristics are shown in Figure 1B. Inclusion criteria were physical inactivity (i.e., inactive in terms of exercise training and job; < 150 min/wk moderate-intensity exercise; no structured physical activity for six months prior to recruitment), no cardiopulmonary abnormalities, no injuries, ability to pass the Exercise and Sport Science Australia (ESSA) pre-exercise screening tool and/or obtain general practitioner clearance to exercise, and ability to ride a stationary cycle at high intensity. Exclusion criteria were known cardiovascular disease or diabetes mellitus, major or chronic illness that impairs mobility and/or eating/digestion, taking prescription medications (i.e., beta-blockers, anti-arrhythmic drugs, statins, insulin sensitizing drugs, or drugs that increase the risk of bleeding [anticoagulants, antiplatelets, novel oral anticoagulants, nonsteroidal anti-inflammatory drugs, selective norepinephrine reuptake inhibitors, or selective serotonin reuptake inhibitors], or known bleeding disorder (i.e., hemophilia A [factor VIII deficiency], von Willebrand disease, or other rare factor deficiencies including I, II, V, VII, X, XI, XII and XIII).

### Participant baseline measurements, exercise testing and familiarization

Participants arrived at the laboratory following a 10-12 h overnight fast for dual-energy X-ray absorptiometry (DXA)-based body composition analysis (Lunar iDXA; GE HealthCare, Chicago, IL, USA). Upon arrival each participant’s height and body mass were recorded, bladder was voided and any metal jewelry or clothing items containing metal were removed prior to DXA scanning. Next, resting metabolic rate (RMR) testing was performed using a calibrated TrueOne 2400 (Parvo Medics, Sandy, UT, USA) with expired air collected for a total of 25 min, including 10 min baseline measurement and 15 min data collection. Following RMR, resting blood pressure and heart rate were recorded in a seated position.

Following baseline measurements, each participant completed an incremental test to volitional fatigue on an electronically braked cycle ergometer (Lode Excalibur Sport; Lode, Groningen, Netherlands) to determine v̇ O_2_peak and PPO. During the maximal exercise capacity test expired gas was collected every 30 s via open-circuit respirometry (TrueOne 2400; Parvo Medics) with continuous heart rate monitoring (Polar Heart Rate Monitor; Polar Electro, Kempele, Finland). Before each test, gas analyzers were calibrated with commercially available gases (16% O_2_, 4% CO_2_), and volume flow was calibrated using a 3L syringe. Following a 5-min warm up at 1 W/kg, resistance was increased by 25 W every 150 s until volitional fatigue, determined as the inability to maintain a cadence > 60 rpm. Individual v̇ O_2_peak and PPO were determined, with PPO calculated as Wfinal + (t/150 * 25) if the final stage was not completed, to calculate the work rate for subsequent work (67.9 ± 10.2 kJ) and duration (10 min) matched HIIT and MICT exercise trials. At least 72 h prior to the first randomized exercise trial, participants returned to the laboratory for exercise trial familiarization. Following a 5-min warmup at 1 W/kg, participants completed two cycling-based exercise sessions consisting of a single bout of HIIT and MICT (10 min total each) to confirm their ability to successfully complete exercise trials at the prescribed intensities.

### Dietary control and standardized meals

Participants recorded dietary information using the Easy Diet Diary mobile phone application. Food and fluid intake for all meals and snacks was recorded over a three-day period and analyzed by an accredited research dietician. The habitual diet record and baseline DXA/RMR data were used to prescribe a standardized meal, which was provided with cooking instructions and consumed at the participant’s home for dinner between 1800-2000 h before each trial day. The macronutrient composition of the standardized dinner was 50% carbohydrate, 30% fat and 20% protein. Participants refrained from consuming any other food or fluids other than water from 2000 h the evening prior to each trial. Participants also refrained from caffeine consumption after 1200 h and alcohol consumption and ibuprofen 24 h prior to each trial.

### Exercise trials and skeletal muscle biopsy collection

In a randomized crossover design, participants were assigned their first exercise trial (i.e., HIIT or MICT, with half of participants randomly assigned to perform the HIIT session first) prior to commencing trial days and did not perform any exercise in the 72 h prior to each trial day. The HIIT session consisted of 10 min total cycling with 1-min intervals at 85 ± 0.1% of individual PPO (176 ± 34 W) interspersed with 1-min active recovery intervals at 50 W. The MICT protocol was work- and duration-matched and consisted of an acute bout of continuous cycling at 55 ± 2% of individual PPO (113 ± 17 W). A schematic of the overall study design is shown in Figure 1A. The two HIIT and MICT exercise trials were separated by ≥ 10 days, and it was not possible to blind participants nor the principal researchers to the order of these trials.

On trial days participants arrived at the laboratory at 0700-0800 h, having fasted overnight since consuming the standardized dinner the evening prior, and only consumed water during the trials. Each participant’s preferred arm was cannulated for blood collections (detailed below) and local anesthetic (1% lignocaine hydrochloride in saline; McFarlane; Surrey Hills, Victoria Australia; 11037-AS) was administered to the vastus lateralis by a highly experienced medical doctor. A percutaneous skeletal muscle biopsy was collected at rest prior to commencing exercise (0 min) using a Bergstrom needle modified with suction and immediately snap-frozen, placed in liquid nitrogen and stored at −80 °C until analysis. Additional skeletal muscle biopsy samples were collected from each participant mid-exercise (5 min) and immediately post-exercise (10 min), with the total 10 min cycling duration consistent for each exercise intensity. For the HIIT trial, the mid-exercise biopsy was collected during an active recovery interval on a bed placed directly behind the cycle ergometer (∼30 sec), and participants re-commenced cycling immediately after the biopsy was collected. For the MICT trial, participants stopped cycling (∼30 sec) for the mid-exercise biopsy collection and re-commenced cycling immediately after the biopsy was collected.

### Blood sampling and analyses

Upon arrival to the laboratory, a cannula (22 G; Terumo, Tokyo, Japan) was inserted into an antecubital vein of each participant. Two vacutainers of venous blood (6 mL each) were collected via cannula at the same timepoints as skeletal muscle biopsies above, including pre-exercise (0 min), mid-exercise (5 min) and immediately post-exercise (10 min). Lipid panels (Roche Diagnostics, Basel, Switzerland; 6380115190) including triglyceride, total cholesterol, high-density lipoproteins (HDL) and low-density lipoproteins (LDL) were immediately measured from an aliquot of whole blood (∼19 μL) using the COBAS b 101 system (Roche Diagnostics). Following inversion ten times, one EDTA-coated vacutainer collected for plasma (Interpath, Somerton, Victoria, Australia; 454036) was immediately placed on ice, centrifuged at 1,500 x *g* for 10 min at 4 °C, aliquoted and stored at −80 °C until analysis. Simultaneous measurement of glucose and lactate from plasma aliquots was performed in duplicate using a calibrated YSI 2900 Biochemistry Analyzer (YSI Incorporated, Yellow Springs, OH, USA).

### Phosphoproteomic sample preparation

As depicted in Figure 2A and detailed below, proteins from each human skeletal muscle biopsy sample were extracted, digested to peptides with trypsin, and isobarically labeled prior to phosphopeptide enrichment, fractionation and analysis by LC-MS/MS. Briefly, ∼30 mg of each snap-frozen human skeletal muscle was lysed as previously described (Blazev et al., 2022) in 6 M guanidine HCL (Sigma, St. Louis, MO, USA; G4505) and 100 mM Tris pH 8.5 containing 10 mM tris(2-carboxyethyl)phosphine (Sigma; 75259) and 40 mM 2-chloroacetamide (Sigma; 22790) using tip-probe sonication. The resulting lysate was heated at 95 °C for 5 min and centrifuged at 20,000 x *g* for 10 min at 4 °C, and the resulting supernatant was diluted 1:1 with water and precipitated with 5 volumes of acetone at −20 °C overnight. Lysate was then centrifuged at 4,000 x *g* for 5 min at 4 °C and the protein pellet was resuspended in Digestion Buffer (10% 2,2,2-Trifluoroethanol (Sigma; 96924) in 100 mM HEPES pH 8.5). Protein content was determined using BCA (Thermo Fisher Scientific, Waltham, MA, USA). Four hundred μg protein (normalized to 100 μL final volume in Digestion Buffer) was digested with sequencing grade trypsin (Sigma; T6567) and LysC (Wako Chemicals, Richmond, VA, USA; 129-02541) at a 1:50 enzyme:substrate ratio at 37 °C overnight with shaking at 2,000 rpm.

Digested peptides were labeled with 800 μg of 10-plex tandem mass tags (TMT) in 50% acetonitrile to a final volume of 200 μL at room temperature for 1.5 h. The TMT reaction was deacylated with 0.3% (w/v) of hydroxylamine for 10 min at room temperature and quenched to final volume of 1% trifluoroacetic acid (TFA). Each experiment consisting of 7 TMT labeled peptides (10 total experiments, each including all 6 timepoints from a single participant’s HIIT and MICT trials and a pooled internal reference mix of peptides consisting of peptides from all 10 participants) was then pooled, resulting in a final amount of 4 mg peptide per TMT 10-plex experiment. The sample identity and labeling channels are uploaded as a table with the raw proteomic data to the ProteomeXchange Consortium via the PRIDE partner repository (see Resource Availability for login details).

Twenty μg of TMT-labeled peptide was removed for total proteome analysis (data not shown, as only 5-10 min of exercise does not affect total muscle protein content), and phosphopeptides were enriched from the remaining digestion of pooled peptides from each experiment using a modified version of the EasyPhos protocol (Humphrey et al., 2018). Briefly, samples were diluted to a final concentration of 50% isopropanol containing 5% TFA and 0.8 mM KH_2_PO_4_. Dilutions were then incubated with 15 mg of TiO_2_ beads (GL Sciences, Tokyo, Japan; 5010-21315) for 8 min at 40 °C with shaking at 2,000 rpm. Beads were washed four times with 60% isopropanol containing 5% TFA and resuspended in 60% isopropanol containing 0.1% TFA. The bead slurry was transferred to in-house packed C8 microcolumns (3M Empore; 11913614) and phosphopeptides were eluted with 40% acetonitrile containing 5% ammonium hydroxide. The enriched phosphopeptides and 20 μg aliquot for total proteome analysis were acidified to a final concentration of 1% TFA in 90% isopropanol and purified by in-house packed SDB-RPS (Sigma; 66886-U) microcolumns. The purified peptides and phosphopeptides were resuspended in 2% acetonitrile in 0.1% TFA and stored at −80 °C prior to offline fractionation using neutral phase C18BEH HPLC as previously described (Blazev et al., 2022).

### LC-MS/MS data acquisition and processing

Peptides were analyzed on a Dionex 3500 nanoHPLC, coupled to an Orbitrap Eclipse mass spectrometer (Thermo Fisher Scientific) via electrospray ionization in positive mode with 1.9 kV at 275 °C and RF set to 30%. Separation was achieved on a 50 cm x 75 μm column (PepSep, Marslev, Denmark) packed with C18-AQ (1.9 μm; Dr Maisch, Ammerbuch, Germany) over 120 min at a flow rate of 300 nL/min. The peptides were eluted over a linear gradient of 3-40% Buffer B (Buffer A: 0.1% formic acid; Buffer B: 80% acetonitrile, 0.1% v/v FA) and the column was maintained at 50 °C. The instrument was operated in data-dependent acquisition (DDA) mode with an MS1 spectrum acquired over the mass range 350-1,550 m/z (120,000 resolution, 1 x 10^6^ automatic gain control (AGC) and 50 ms maximum injection time) followed by MS/MS analysis with fixed cycle time of 3 s via HCD fragmentation mode and detection in the orbitrap (50,000 resolution, 1 x 10^5^ AGC, 150 ms maximum injection time, and 0.7 m/z isolation width). Only ions with charge state 2-7 triggered MS/MS with peptide monoisotopic precursor selection and dynamic exclusion enabled for 30 s at 10 ppm.

DDA data were searched against the UniProt human database (June 2020; UP000005640_9606 and UP000005640_9606_additional) with MaxQuant v1.6.7.0 using default parameters with peptide spectral matches, peptide and protein FDR set to 1% (Cox and Mann, 2008). All data were searched with oxidation of methionine set as a variable modification and cysteine carbamidomethylation set as a fixed modification. For analysis of phosphopeptides, phosphorylation of Serine, Threonine and Tyrosine was set as a variable modification, and for analysis of TMT-labeled peptides, TMT was added as a fixed modification to peptide N-termini and Lysine. First search MS1 mass tolerance was set to 20 ppm followed by recalibration and main search MS1 tolerance set to 4.5 ppm, while MS/MS mass tolerance was set to 20 ppm. MaxQuant output data were initially processed with Perseus (Tyanova et al., 2016) to remove decoy data, potential contaminants and proteins only identified with a single peptide containing oxidized methionine. The ‘Expand Site’ function was additionally used for phosphoproteomic data to account for multi-phosphorylated peptides prior to statistical analysis.

### Bioinformatic analysis

For analysis of human muscle phosphopeptides with TMT-based quantification, data were first log_2_ transformed and each phosphosite abundance was corrected by subtracting the abundance of the pooled sample in the same TMT batch. The phosphoproteomic data were processed using the pipeline implemented in the PhosR package (Kim et al., 2021a). Filtering was performed to retain phosphosites present in at least three participants (out of ten total participants), in at least one timepoint (out of six total timepoints). Missing values in the retained phosphosites were imputed first by a site- and sample condition-specific imputation method, where for a phosphosite that contains missing values in a condition, if more than three samples were quantified in that condition, the missing values were imputed based on these quantified values for that phosphosite in that condition, and then by a random-tail imputation method (Kim et al., 2021b). The imputed data were normalized using ‘Combat’ function in sva package (Johnson et al., 2007) for removing batch effects and then ‘RUVphospho’ function in PhosR for the removal of additional unwanted variation with a set of stably phosphorylated sites as negative controls (Xiao et al., 2022). The batch-corrected data were further converted to ratios relative to the pre-exercise samples (i.e., ‘0 min’ controls).

Differentially phosphorylated sites were identified using limma R package (Ritchie et al., 2015). Phosphosites with −/+ 1.5-fold change and FDR-adjusted *P* value < 0.05 were considered as differentially phosphorylated (**Figure 3A-H**). Kinase activities at post-or mid-exercise for both HIIT and MICT were inferred based on the changes in phosphorylation (relative to the corresponding pre-exercise control samples) of their known substrates using the KinasePA package (Yang et al., 2016) and the PhosphoSitePlus annotation database (Hornbeck et al., 2012) (**Figure 4A**). Pathway enrichment analysis was then performed, whereby phosphosites were first summarised into their host protein levels by taking the maximum log_2_ fold change for each comparison between conditions, and then the pathway enrichment was performed based on the inferred host protein changes using the KinasePA package and Reactome database (Joshi-Tope et al., 2005) (**Figure 4B**).

Putative substrates of kinases for HIIT and MICT were predicted using the ‘kinaseSubstrateScore’ function in the PhosR package, and the results were represented as heatmaps (**Figure 5A-B**). Pathway over-representation analysis was performed on protein sets identified from the kinase-substrate scoring analysis (i.e., kinase-substrate pairs with score > 0.85 were selected) per kinase using the ‘enrichKEGG‘ function implemented in the clusterProfiler R package (Yu et al., 2012), and *P* values were adjusted for multiple testing using Benjamini Hochberg FDR correction at *α* = 0.05 (**Figure 5C-D**). The prediction scores were subsequently used for constructing signalome networks. Pearson’s correlation was performed on pairwise kinases, and then the correlation matrix was binarized based on the correlation score threshold of 0.85. Undirected graphs were built from the binary adjacency matrix using the ‘graph_from_adjacency_matrix’ function from the igraph package (Csárdi G, 2024), and results from this analysis were presented as network diagrams (**Figure 6A-B**).

Five protein modules were identified by clustering proteins with phosphosites sharing similar dynamic phosphorylation profiles and kinase regulation across both HIIT and MICT. The proportion of phosphosites that were phosphorylated by kinases for each protein module was calculated and presented as bubble plots (**Figure 6C-D**). The activity of each protein module was then inferred. The regulated phosphosites were first obtained across all conditions (i.e., ANOVA test with adjusted *P* < 0.05), and the average log_2_ fold change of the regulated phosphosites for each of the five modules (relative to the corresponding pre-exercise control) were calculated (**Supplemental Figure 1A-E**).

### Statistical analysis

Statistical analysis of plasma lactate and glucose concentrations was performed using GraphPad Prism (version 9.4). A two-way ANOVA with repeated measurements was used to determine the effects of time and exercise intensity, with Tukey’s test applied for multiple comparisons (*P* < 0.05 considered as significant; sample size and statistical parameters reported in the **Figure 1** legend). Spearman correlation of individual phosphorylation sites with plasma lactate concentrations at each timepoint and exercise intensity was performed to determine significantly correlated phosphosites, with Benjamini Hochberg false discovery rate applied (q < 0.05 considered as significant; sample size and statistical parameters reported in the **Figure 7** legend).

## REFERENCES

Bartlett, J.D., Hwa Joo, C., Jeong, T.S., Louhelainen, J., Cochran, A.J., Gibala, M.J., Gregson, W., Close, G.L., Drust, B., and Morton, J.P. (2012). Matched work high-intensity interval and continuous running induce similar increases in PGC-1alpha mRNA, AMPK, p38, and p53 phosphorylation in human skeletal muscle. J Appl Physiol (1985) 112, 1135–1143.

Bishop, D.J., Botella, J., and Granata, C. (2019). CrossTalk opposing view: Exercise training volume is more important than training intensity to promote increases in mitochondrial content. J Physiol 597, 4115–4118.

Blazev, R., Carl, C.S., Ng, Y.K., Molendijk, J., Voldstedlund, C.T., Zhao, Y., Xiao, D., Kueh, A.J., Miotto, P.M., Haynes, V.R., et al. (2022). Phosphoproteomics of three exercise modalities identifies canonical signaling and C18ORF25 as an AMPK substrate regulating skeletal muscle function. Cell Metab 34, 1561–1577 e1569.

Brooks, G.A., Osmond, A.D., Arevalo, J.A., Duong, J.J., Curl, C.C., Moreno-Santillan, D.D., and Leija, R.G. (2023). Lactate as a myokine and exerkine: drivers and signals of physiology and metabolism. J Appl Physiol (1985) 134, 529-548.

Campbell, W.W., Kraus, W.E., Powell, K.E., Haskell, W.L., Janz, K.F., Jakicic, J.M., Troiano, R.P., Sprow, K., Torres, A., Piercy, K.L., et al. (2019). High-Intensity Interval Training for Cardiometabolic Disease Prevention. Med Sci Sports Exerc 51, 1220–1226.

Cartee, G.D. (2015). Roles of TBC1D1 and TBC1D4 in insulin- and exercise-stimulated glucose transport of skeletal muscle. Diabetologia 58, 19–30.

Cochran, A.J., Percival, M.E., Tricarico, S., Little, J.P., Cermak, N., Gillen, J.B., Tarnopolsky, M.A., and Gibala, M.J. (2014). Intermittent and continuous high-intensity exercise training induce similar acute but different chronic muscle adaptations. Exp Physiol 99, 782–791.

Combes, A., Dekerle, J., Webborn, N., Watt, P., Bougault, V., and Daussin, F.N. (2015). Exercise-induced metabolic fluctuations influence AMPK, p38-MAPK and CaMKII phosphorylation in human skeletal muscle. Physiol Rep 3.

Cox, J., and Mann, M. (2008). MaxQuant enables high peptide identification rates, individualized p.p.b.-range mass accuracies and proteome-wide protein quantification. Nat Biotechnol 26, 1367–1372.

Csárdi G, N.T., Traag V, Horvát S, Zanini F, Noom D, Müller K (2024). igraph: Network Analysis and Visualization in R.

Ducommun, S., Deak, M., Zeigerer, A., Goransson, O., Seitz, S., Collodet, C., Madsen, A.B., Jensen, T.E., Viollet, B., Foretz, M., et al. (2019). Chemical genetic screen identifies Gapex-5/GAPVD1 and STBD1 as novel AMPK substrates. Cell Signal 57, 45–57.

Egan, B., and Zierath, J.R. (2013). Exercise metabolism and the molecular regulation of skeletal muscle adaptation. Cell Metab 17, 162–184.

Fiorenza, M., Gunnarsson, T.P., Hostrup, M., Iaia, F.M., Schena, F., Pilegaard, H., and Bangsbo, J. (2018). Metabolic stress-dependent regulation of the mitochondrial biogenic molecular response to high-intensity exercise in human skeletal muscle. J Physiol 596, 2823–2840.

Furrer, R., Hawley, J.A., and Handschin, C. (2023a). The molecular athlete: exercise physiology from mechanisms to medals. Physiol Rev 103, 1693–1787.

Furrer, R., Heim, B., Schmid, S., Dilbaz, S., Adak, V., Nordstrom, K.J.V., Ritz, D., Steurer, S.A., Walter, J., and Handschin, C. (2023b). Molecular control of endurance training adaptation in male mouse skeletal muscle. Nat Metab 5, 2020–2035.

Gibala, M.J., and Hawley, J.A. (2017). Sprinting Toward Fitness. Cell Metab 25, 988–990.

Gibala, M.J., Little, J.P., Macdonald, M.J., and Hawley, J.A. (2012). Physiological adaptations to low-volume, high-intensity interval training in health and disease. J Physiol 590, 1077–1084.

Gibala, M.J., McGee, S.L., Garnham, A.P., Howlett, K.F., Snow, R.J., and Hargreaves, M. (2009). Brief intense interval exercise activates AMPK and p38 MAPK signaling and increases the expression of PGC-1alpha in human skeletal muscle. J Appl Physiol (1985) 106, 929–934.

Hargreaves, M., and Spriet, L.L. (2020). Skeletal muscle energy metabolism during exercise. Nat Metab 2, 817–828.

Hawley, J.A., Hargreaves, M., Joyner, M.J., and Zierath, J.R. (2014). Integrative biology of exercise. Cell 159, 738–749.

Hawley, J.A., Lundby, C., Cotter, J.D., and Burke, L.M. (2018). Maximizing Cellular Adaptation to Endurance Exercise in Skeletal Muscle. Cell Metab 27, 962–976.

Hoffman, N.J. (2017). Omics and Exercise: Global Approaches for Mapping Exercise Biological Networks. Cold Spring Harb Perspect Med 7.

Hoffman, N.J., Parker, B.L., Chaudhuri, R., Fisher-Wellman, K.H., Kleinert, M., Humphrey, S.J., Yang, P., Holliday, M., Trefely, S., Fazakerley, D.J., et al. (2015). Global Phosphoproteomic Analysis of Human Skeletal Muscle Reveals a Network of Exercise-Regulated Kinases and AMPK Substrates. Cell Metab 22, 922–935.

Hornbeck, P.V., Kornhauser, J.M., Tkachev, S., Zhang, B., Skrzypek, E., Murray, B., Latham, V., and Sullivan, M. (2012). PhosphoSitePlus: a comprehensive resource for investigating the structure and function of experimentally determined post-translational modifications in man and mouse. Nucleic Acids Res 40, D261–270.

Hostrup, M., Lemminger, A.K., Stocks, B., Gonzalez-Franquesa, A., Larsen, J.K., Quesada, J.P., Thomassen, M., Weinert, B.T., Bangsbo, J., and Deshmukh, A.S. (2022). High-intensity interval training remodels the proteome and acetylome of human skeletal muscle. Elife 11.

Humphrey, S.J., Karayel, O., James, D.E., and Mann, M. (2018). High-throughput and high-sensitivity phosphoproteomics with the EasyPhos platform. Nat Protoc 13, 1897–1916.

Johnson, W.E., Li, C., and Rabinovic, A. (2007). Adjusting batch effects in microarray expression data using empirical Bayes methods. Biostatistics 8, 118–127.

Joshi-Tope, G., Gillespie, M., Vastrik, I., D’Eustachio, P., Schmidt, E., de Bono, B., Jassal, B., Gopinath, G.R., Wu, G.R., Matthews, L., et al. (2005). Reactome: a knowledgebase of biological pathways. Nucleic Acids Res 33, D428–432.

Kim, H.J., Kim, T., Hoffman, N.J., Xiao, D., James, D.E., Humphrey, S.J., and Yang, P. (2021a). PhosR enables processing and functional analysis of phosphoproteomic data. Cell Rep 34, 108771.

Kim, H.J., Kim, T., Xiao, D., and Yang, P. (2021b). Protocol for the processing and downstream analysis of phosphoproteomic data with PhosR. STAR Protoc 2, 100585.

Kristensen, D.E., Albers, P.H., Prats, C., Baba, O., Birk, J.B., and Wojtaszewski, J.F. (2015). Human muscle fibre type-specific regulation of AMPK and downstream targets by exercise. J Physiol 593, 2053–2069.

Little, J.P., Safdar, A., Bishop, D., Tarnopolsky, M.A., and Gibala, M.J. (2011). An acute bout of high-intensity interval training increases the nuclear abundance of PGC-1alpha and activates mitochondrial biogenesis in human skeletal muscle. Am J Physiol Regul Integr Comp Physiol 300, R1303–1310.

MacInnis, M.J., and Gibala, M.J. (2017). Physiological adaptations to interval training and the role of exercise intensity. J Physiol 595, 2915–2930.

MacInnis, M.J., Skelly, L.E., and Gibala, M.J. (2019). CrossTalk proposal: Exercise training intensity is more important than volume to promote increases in human skeletal muscle mitochondrial content. J Physiol 597, 4111–4113.

MacInnis, M.J., Zacharewicz, E., Martin, B.J., Haikalis, M.E., Skelly, L.E., Tarnopolsky, M.A., Murphy, R.M., and Gibala, M.J. (2017). Superior mitochondrial adaptations in human skeletal muscle after interval compared to continuous single-leg cycling matched for total work. J Physiol 595, 2955–2968.

McConell, G.K., Lee-Young, R.S., Chen, Z.P., Stepto, N.K., Huynh, N.N., Stephens, T.J., Canny, B.J., and Kemp, B.E. (2005). Short-term exercise training in humans reduces AMPK signalling during prolonged exercise independent of muscle glycogen. J Physiol 568, 665–676.

Milanovic, Z., Sporis, G., and Weston, M. (2015). Effectiveness of High-Intensity Interval Training (HIT) and Continuous Endurance Training for VO2max Improvements: A Systematic Review and Meta-Analysis of Controlled Trials. Sports Med 45, 1469–1481.

Morita, M., Prudent, J., Basu, K., Goyon, V., Katsumura, S., Hulea, L., Pearl, D., Siddiqui, N., Strack, S., McGuirk, S., et al. (2017). mTOR Controls Mitochondrial Dynamics and Cell Survival via MTFP1. Mol Cell 67, 922–935 e925.

Nsiah-Sefaa, A., Brown, E.L., Russell, A.P., and Foletta, V.C. (2014). New gene targets of PGC-1alpha and ERRalpha co-regulation in C2C12 myotubes. Mol Biol Rep 41, 8009–8017.

Patitucci, C., Hernandez-Camacho, J.D., Vimont, E., Yde, S., Cokelaer, T., Chaze, T., Giai Gianetto, Q., Matondo, M., Gazi, A., Nemazanyy, I., et al. (2023). Mtfp1 ablation enhances mitochondrial respiration and protects against hepatic steatosis. Nat Commun 14, 8474.

Peake, J.M., Tan, S.J., Markworth, J.F., Broadbent, J.A., Skinner, T.L., and Cameron-Smith, D. (2014). Metabolic and hormonal responses to isoenergetic high-intensity interval exercise and continuous moderate-intensity exercise. Am J Physiol Endocrinol Metab 307, E539–552.

Perez-Riverol, Y., Bai, J., Bandla, C., Garcia-Seisdedos, D., Hewapathirana, S., Kamatchinathan, S., Kundu, D.J., Prakash, A., Frericks-Zipper, A., Eisenacher, M., et al. (2022). The PRIDE database resources in 2022: a hub for mass spectrometry-based proteomics evidences. Nucleic Acids Res 50, D543–D552.

Perrini, S., Henriksson, J., Zierath, J.R., and Widegren, U. (2004). Exercise-induced protein kinase C isoform-specific activation in human skeletal muscle. Diabetes 53, 21–24.

Perry, C.G., Lally, J., Holloway, G.P., Heigenhauser, G.J., Bonen, A., and Spriet, L.L. (2010). Repeated transient mRNA bursts precede increases in transcriptional and mitochondrial proteins during training in human skeletal muscle. J Physiol 588, 4795–4810.

Pileggi, C.A., Hedges, C.P., D’Souza, R.F., Durainayagam, B.R., Zeng, N., Figueiredo, V.C., Hickey, A.J.R., Mitchell, C.J., and Cameron-Smith, D. (2023). Minimal adaptation of the molecular regulators of mitochondrial dynamics in response to unilateral limb immobilisation and retraining in middle-aged men. Eur J Appl Physiol 123, 249–260.

Reisman, E.G., Hawley, J.A., and Hoffman, N.J. (2024). Exercise-Regulated Mitochondrial and Nuclear Signalling Networks in Skeletal Muscle. Sports Med.

Richter, E.A., Vistisen, B., Maarbjerg, S.J., Sajan, M., Farese, R.V., and Kiens, B. (2004). Differential effect of bicycling exercise intensity on activity and phosphorylation of atypical protein kinase C and extracellular signal-regulated protein kinase in skeletal muscle. J Physiol 560, 909–918.

Ritchie, M.E., Phipson, B., Wu, D., Hu, Y., Law, C.W., Shi, W., and Smyth, G.K. (2015). limma powers differential expression analyses for RNA-sequencing and microarray studies. Nucleic Acids Res 43, e47.

Robinson, M.M., Dasari, S., Konopka, A.R., Johnson, M.L., Manjunatha, S., Esponda, R.R., Carter, R.E., Lanza, I.R., and Nair, K.S. (2017). Enhanced Protein Translation Underlies Improved Metabolic and Physical Adaptations to Different Exercise Training Modes in Young and Old Humans. Cell Metab 25, 581–592.

Rose, A.J., and Hargreaves, M. (2003). Exercise increases Ca2+-calmodulin-dependent protein kinase II activity in human skeletal muscle. J Physiol 553, 303–309.

Rose, A.J., Kiens, B., and Richter, E.A. (2006). Ca2+-calmodulin-dependent protein kinase expression and signalling in skeletal muscle during exercise. J Physiol 574, 889–903.

Rose, A.J., Michell, B.J., Kemp, B.E., and Hargreaves, M. (2004). Effect of exercise on protein kinase C activity and localization in human skeletal muscle. J Physiol 561, 861–870.

Sollanek, K.J., Burniston, J.G., Kavazis, A.N., Morton, A.B., Wiggs, M.P., Ahn, B., Smuder, A.J., and Powers, S.K. (2017). Global Proteome Changes in the Rat Diaphragm Induced by Endurance Exercise Training. PLoS One 12, e0171007.

Tilokani, L., Russell, F.M., Hamilton, S., Virga, D.M., Segawa, M., Paupe, V., Gruszczyk, A.V., Protasoni, M., Tabara, L.C., Johnson, M., et al. (2022). AMPK-dependent phosphorylation of MTFR1L regulates mitochondrial morphology. Sci Adv 8, eabo7956.

Torma, F., Gombos, Z., Jokai, M., Takeda, M., Mimura, T., and Radak, Z. (2019). High intensity interval training and molecular adaptive response of skeletal muscle. Sports Med Health Sci 1, 24–32.

Trewin, A.J., Parker, L., Shaw, C.S., Hiam, D.S., Garnham, A., Levinger, I., McConell, G.K., and Stepto, N.K. (2018). Acute HIIE elicits similar changes in human skeletal muscle mitochondrial H(2)O(2) release, respiration, and cell signaling as endurance exercise even with less work. Am J Physiol Regul Integr Comp Physiol 315, R1003–R1016.

Tyanova, S., Temu, T., Sinitcyn, P., Carlson, A., Hein, M.Y., Geiger, T., Mann, M., and Cox, J. (2016). The Perseus computational platform for comprehensive analysis of (prote)omics data. Nat Methods 13, 731–740.

Xiao, D., Kim, H.J., Pang, I., and Yang, P. (2022). Functional analysis of the stable phosphoproteome reveals cancer vulnerabilities. Bioinformatics 38, 1956–1963.

Yang, P., Patrick, E., Humphrey, S.J., Ghazanfar, S., James, D.E., Jothi, R., and Yang, J.Y. (2016). KinasePA: Phosphoproteomics data annotation using hypothesis driven kinase perturbation analysis. Proteomics 16, 1868–1871.

Yu, G., Wang, L.G., Han, Y., and He, Q.Y. (2012). clusterProfiler: an R package for comparing biological themes among gene clusters. OMICS 16, 284–287.

Yu, M., Stepto, N.K., Chibalin, A.V., Fryer, L.G., Carling, D., Krook, A., Hawley, J.A., and Zierath, J.R. (2003). Metabolic and mitogenic signal transduction in human skeletal muscle after intense cycling exercise. J Physiol 546, 327–335.

