## Supplementary figures and images for "Phosphoproteomics uncovers exercise intensity-specific signaling networks underlying high-intensity interval training in human skeletal muscle"

### Supplemental Figure 1

#### Supplemental Figure 1

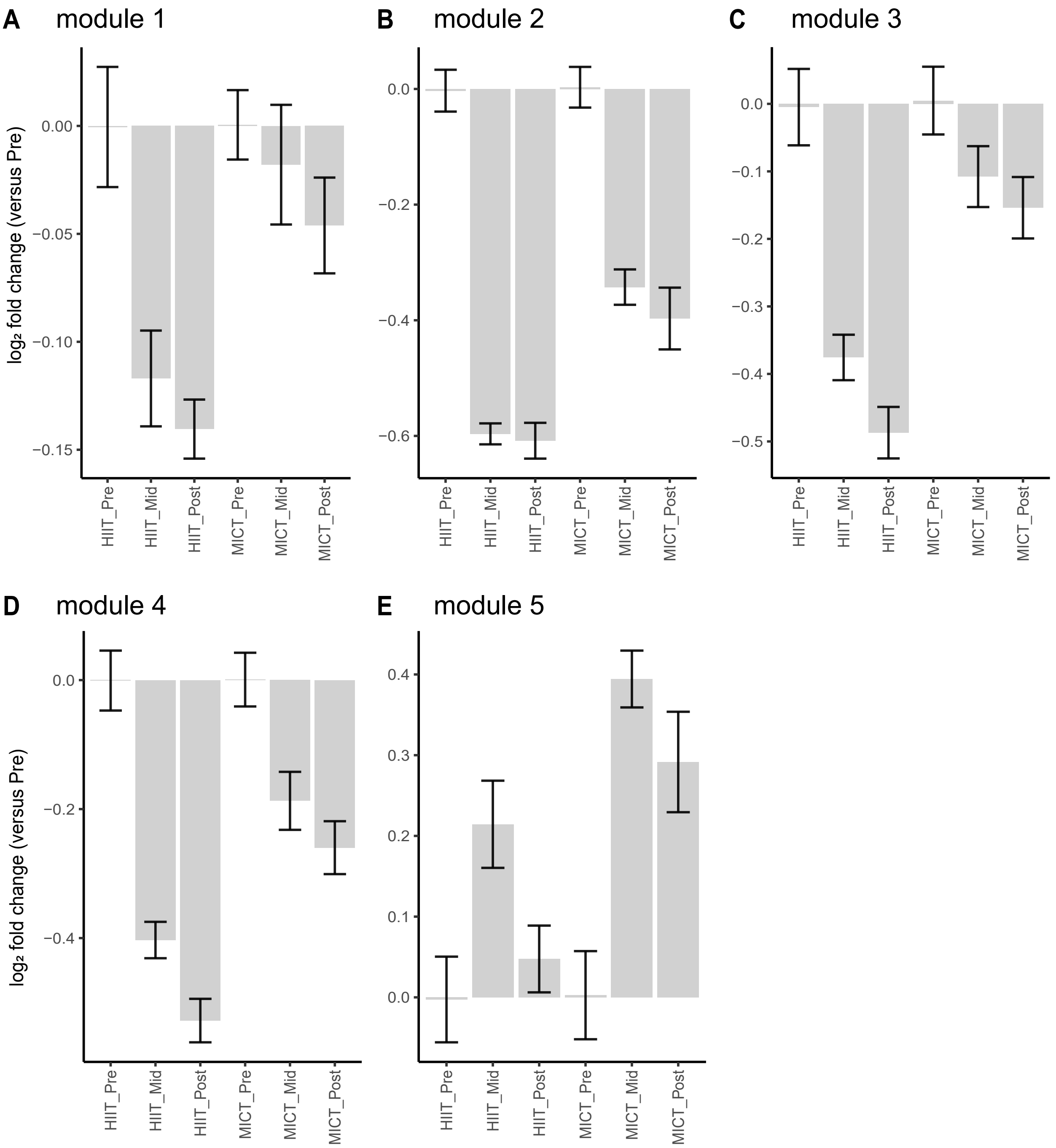
